# Inequalities in childhood pneumococcal conjugate vaccine uptake in England before and after the change from a 2+1 to 1+1 schedule: a longitudinal study

**DOI:** 10.64898/2025.12.05.25341691

**Authors:** Praise Ilechukwu, Daniel Hungerford, Neil French, Edward M. Hill

## Abstract

**Background:** Introducing the pneumococcal conjugate vaccines (PCV) into the routine childhood immunisation schedule in England has reduced pneumococcal disease burden. Nonetheless, pneumococcal disease continues to a]lict marked morbidity and mortality in the population, including young children. In January 2020, England transitioned from a “2+1” PCV schedule (two primary doses at 8 and 16 weeks, with a booster dose at 12 months) to a “1+1” PCV schedule (single primary dose at 12 weeks and a booster dose at 12 months). While immunogenicity studies suggested comparable protection, reducing primary vaccine doses places greater emphasis on timely booster uptake.

**Methods:** We examine national trends in PCV uptake before and after the schedule change and quantify inequalities by deprivation. We analysed quarterly vaccine uptake data for 2013-2025 from the Cover of Vaccination Evaluated Rapidly (COVER) programme for upper-tier local authorities in England linked to 2019 Index of Multiple Deprivation quintiles. We examined booster gaps - the di]erence between primary coverage at 12 months and booster coverage at 24 months. We estimated susceptibility to pneumococcal disease (by birth cohort stratified by quarter) by combining observed PCV uptake with published vaccine effectiveness estimates.

**Findings:** Booster retention deteriorated following the schedule change; mean booster gaps increased from 2.32% (2+1 period) to 4.79% (1+1 period). The largest booster gaps arose in London boroughs. The gap between least and most deprived quintiles widened from 2-3% (2+1 period) to 4-6% (1+1 period). Susceptibility calculations returned notable geographical variation in estimated vaccine type invasive pneumococcal disease susceptibility in England, with disproportionate burden in deprived areas.

**Interpretations:** The success of the 1+1 schedule depends on maintaining equitable, high booster uptake. Declining booster retention and widening inequalities threaten programme e]ectiveness. Increased resource allocation for child immunisations, strengthened follow-up systems and targeted interventions in deprived communities are essential to prevent widening protection gaps.

**Funding:** No specific funding for this project.

**Research in context:** *Evidence before this study:* Pneumococcal conjugate vaccines (PCVs) have been part of the routine childhood immunisation schedule in England since September 2006, firstly using PCV7 (targeting seven serotypes) and then from 2010 PCV13 (targeting thirteen serotypes). This has resulted in reduced invasive pneumococcal disease (IPD) burden, although IPD continues to cause marked morbidity and mortality across the population, including young children. Originally administered as a “2+1” schedule (two primary doses at 8 and 16 weeks, with a booster dose at 12 months), England transitioned In January 2020 to a “1+1” PCV schedule (single primary dose at 12 weeks and a booster dose at 12 months). While immunogenicity studies suggested comparable protection, reducing primary vaccine doses places greater emphasis on timely booster uptake. We searched PubMed, Google Scholar, and medRxiv for articles published in English from inception to 30 Nov 2025 with the following search terms: (“PCV-13” OR “13-valent pneumococcal conjugate vaccine” OR “pneumococcal conjugate vaccine”) AND (booster OR “third dose” OR “2+1 schedule” OR “additional dose”) AND (children OR infants OR toddlers OR paediatric OR paediatric) AND (“vaccine effectiveness” OR “vaccine efficacy”) AND “England”. To our knowledge, an observational analysis investigating potential inequalities in Childhood Pneumococcal Conjugate Vaccine Uptake and resultant susceptibility to IPD in England Before and After the change from a 2+1 to 1+1 schedule in January 2020 has not been conducted.

*Added value of this study:* We analysed temporal and spatial trends and inequalities in PCV13 uptake across England at an Upper Tier Local Authority level for 2013-2025. This period included the January 2020 transition from a 2+1 to 1+1 dose schedule. The analysis revealed three critical findings: booster retention deteriorated under the 1+1 schedule; socioeconomic inequalities in vaccine uptake persisted and widened (particularly affecting the most deprived communities); using a susceptibility calculation that combined PCV uptake data with current knowledge on vaccine effectiveness estimates for PCV13 against vaccine type IPD, we highlight a growing inequitable susceptibility to vaccine type IPD in child cohorts.

*Implications of all the available evidence:* Our study shows that PCV booster retention has notably declined in England since the change from a ‘2+1’ to a ‘1+1’ schedule change. This means the full protective potential of the 1+1 schedule is not being realised. A trend of lower booster retention amongst children in more deprived areas risks avoidable disease burden being concentrated in the most disadvantaged communities and widening health inequalities. Other countries considering a ‘1+1’ schedule change should consider underlying inequality in vaccine uptake and booster retention before implementation. Systems strengthening and targeted, equity-focused interventions are needed to address the identified coverage gaps.

## 1. Introduction

Pneumococcal disease, caused by *Streptococcus pneumoniae*, remains a leading cause of vaccine-preventable illness and death globally, particularly in children under five years old. In 2019, globally an estimated 740,180 deaths (14% of all deaths) in children under five were attributable to S. pneumoniae ^1^. In England, the introduction of pneumococcal conjugate vaccines (PCVs) into the routine childhood immunisation schedule has led to declines in invasive pneumococcal disease (IPD) through direct and indirect (herd) immunity. PCV7 (targeting seven serotypes) was first introduced in September 2006, followed by PCV13 (targeting thirteen serotypes) in 2010 ^2^.

Within four years of both programmes, the maximum benefit of reducing IPD vaccine type (VT) serotypes - was reached, with a plateauing of VT IPD cases across all age groups (contrasting with global-level trends of plateauing VT IPD rates in children under 5 years of age seven years after introduction of higher-valency PCVs, and continued decline in VT IPD rates in older children and adults up to nine years after introduction of higher-valency PCVs ^3^). Prior to 2020, England’s infant vaccination programme followed a 2+1 PCV13 schedule: two primary doses at 8 and 16 weeks, followed by a booster at 12 months. In January 2020 the UK became the first and to date, only European nation to have a reduced 1+1 schedule, a primary dose at 12 weeks and a booster at 12-months ^4^. The rationale for was supported by clinical trials and dynamic transmission modelling, which suggested similar levels of population protection could be maintained, provided that the booster uptake remained high ^5^.

Following the change in schedule, overall IPD incidence in 2022-23 remained 14% lower than pre-COVID19 pandemic levels in 2019-20 (4598 cases vs 5316 cases; IRR 0.86, 95% CI 0.81-0.91; p<0.001), though incidence was 34% higher in children aged <15 years (378 cases vs 292 cases; IRR 1.34, 95% CI 1.08-1.68; p=0.009). This time also saw society-wide public health and social measures (PHSMs) used in response to the COVID-19 pandemic, which dramatically reduced the incidence of contagious infectious diseases including IPD. How patterns of infectious diseases will respond to the perturbed immune landscape remains unclear ^6^.

Importantly, breakthrough infections and vaccine failure rates were not significantly different between 1+1 eligible children in 2022–23 compared with 2+1 eligible children between 2017–18 and 2019–20 (1.08 per 100,00 person-years [19 of 1,758,189 livebirths] vs 0.76 per 100,000 person-years [44 of 5,792,902 livebirths]; IRR 1.42, 95% CI 0.78-2.49; p=0.20) ^2^. These observations should be considered in the context of COVID-19 pandemic PHSMs resulting in the sustained suppression of multiple vaccine-preventable infectious diseases after the removal of restrictions in England ^7^.

Despite these assurances, the adequacy of individual-level protection under the reduced schedule is highly dependent on maintaining high vaccine uptake of the booster dose. A recent study showed that uptake of paediatric vaccines in England, have declined since 2019 and inequalities in uptake rates were widening^8^. National PCV booster coverage at 24 months has declined steadily since its peak of 92.5% in 2012–13, falling to 88.2% in 2023–24 ^9^. These trends raise questions about the suitability and effectiveness of the 1+1 schedule against a backdrop of declining paediatric vaccine coverage, particularly in the context of existing health inequalities.

In England, vaccine uptake is lower in socioeconomic disadvantaged populations and there is a clear social gradient linked to deprivation for IPD burden ^8^. National surveillance data from the Northeast of England from April 2006 to March 2011 show that the incidence of IPD increased linearly from 7.0 cases per 100,000 population in the least socioeconomically deprived quintile to 13.6 cases per 100,000 in the most deprived ^10^.

This paper explores two key areas of public health interest in the context of this schedule change. First, we investigated the temporal and spatial trends and inequalities in paediatric PCV13 uptake across cohorts receiving the 2+1 versus the 1+1 schedule in England. Second, we estimated and compared pneumococcal disease susceptibility among different cohorts; susceptibility in this context refers to the proportion of children in the population at risk to VT IPD – IPD caused by serotypes covered by PCV13 - due to either non-vaccination, partial vaccination (i.e. not completing the full schedule of doses) plus the residual risk of disease after full vaccination. By inferring susceptibility at different time points using vaccine effectiveness estimates against IPD available from the literature, we draw conclusions on how variations in booster uptake might influence susceptibility to VT IPD in early childhood.

## 2. Methods

Herein we overview our methodology to process the PCV uptake date from COVER (Section 2.1.1) and its linkage to a validated measure of socioeconomic deprivation for small areas in England, the Index of Multiple Deprivation (Section 2.1.2); analyse the PCV uptake data (Section 2.2); estimate pneumococcal disease susceptibility (Section 2.3).

We performed all data manipulation, statistical analysis and visualisation using R version 4.4.1^11^. All analysis code and cleaned datasets are publicly available: https://github.com/aaliswalker/DASC500-Dissertation---Inequalities-in-Childhood-Pneumococcal-Vaccine-Uptake-in-England

### 2.1 Datasets

#### 2.1.1. PCV uptake data from COVER

We extracted quarterly PCV uptake data for England from the UK Health Security Agency (UKHSA) Cover of Vaccination Evaluated Rapidly (COVER) programme ^12^. These data spanned Q2 2013-2014 to Q4 2024–2025, where quarters represent financial year periods: Q1 April-June, Q2 July-September, Q3 October-December, Q4 January-March (using 2024-2025 as an example, Q1 2024-2025 corresponds to April-June 2024, Q2 2024-2025 corresponds to July-September 2024, Q3 2024-2025 corresponds to October-December 2024 and Q4 2025-25 corresponds to January-March 2025). The data reported PCV uptake at 12 months (primary course) and 24 months (booster course) per upper-tier local authority (UTLA); a UTLA is a geographical area type in England – as of 2023 there were 153 UTLAs in England – and are responsible for local services ^13^. Denominator eligible population counts for each quarter and UTLA corresponded to: for the 12-month primary dose cohorts, children in that UTLA who reached 12 months of age during that quarter; for the 24-month booster dose cohorts, children in that UTLA who reached 24 months of age during that quarter. The start and end of the COVID-19 period (11 March 2020 - 5 May 2023) were informed by available NHS data ^14^. See the S1 Text for additional information on COVER dataset processing.

#### 2.1.2. Linkage of vaccine uptake data to Index of Multiple Deprivation

The COVER data contained Office for National Statistics (ONS) UTLA codes. We therefore used the ONS UTLA codes to link the COVER vaccine uptake data to the 2019 Index of Multiple Deprivation (IMD) for England ^15^.

Geographically, with initial consideration of all areas of England, we excluded the Isles of Scilly of The City of London from all analyses due to their very small populations (less than 10,000 in the 2021 census) and unique administrative arrangements ^16^. For clarity, The City of London is a distinct local authority with a very small resident population and differs from the Greater London area, which refers to 32 London UTLAs (also referred to as boroughs) with a population of over 9 million. Not all UTLAs appeared in every quarterly COVER file due to administrative boundary changes during the study period. We identified and resolved several additional UTLA code inconsistencies (see S2 Text for further details). The final analytical dataset comprised 6,816 observations across 149 distinct UTLAs.

IMD is the official measure of relative deprivation for Lower-Layer Super Output Areas (LSOAs) in England. There are over 30,000 LSOAs in England, which are a standard statistical geography designed to be of a similar population size, with an average of approximately 1,500 residents or 650 households. The IMD ranks the LSOAs in England by producing an overall relative measure of deprivation by combining information from seven different domains of deprivation: income deprivation, employment deprivation, education, skills and training deprivation, health deprivation and disability, crime, barriers to housing and services, living environment deprivation ^15^. We acknowledge that we treated IMD as a static measure, whereas it can have temporal variation. That being said, analysis of the IMD from 2004 to 2015 found overall and health-related deprivation patterns to have persisted in England, with large and unchanging health inequalities between the North and the South ^17^.

We used the IMD average score for each UTLA as reported in the 2019 Index of Multiple Deprivation for England ^15^. The IMD average score is a population weighted average of the combined scores for the LSOAs in a larger area (calculated by averaging the LSOA scores in each larger area after they have been population weighted); note that highly polarised areas in terms of deprivation tend to score higher on the average score measure compared to an average rank measure. Following an approach used in recent studies examining local government inequalities (e.g. Murrell et al. (2024) ^18^), we applied quintile-based stratification to the population-weighted UTLA IMD average scores (‘quintile 1’ corresponded to the least deprived areas; ‘quintile 5’ corresponded to the most deprived areas). See S3 Text for a listing of UTLAs in each IMD quintile.

### 2.2. Analysis of PCV Uptake Data

#### 2.2.1. Temporal trends in PCV uptake

We analysed national PCV uptake trends using weighted means (weighted by population denominators to account for varying local authority sizes). We inspected time series trends in 12-month primary coverage and 24-month booster coverage across the study period by IMD. We also examined spatial patterns through choropleth mapping linking UTLA boundaries to uptake data via ONS codes ^19^.

#### 2.2.2. Booster retention analysis

We next examined changes in booster retention patterns following the schedule transition. For each quarter and each of the retained UTLAs, we computed the percentage point difference between the mean 12-month primary coverage one year prior and the mean 24-month booster coverage at the reference quarter. As an example, to examine booster retention for Q1 2023 we used the mean 12-month primary coverage from Q1 2022 and the mean 24-month booster coverage from Q1 2023.

We stratified our analysis by schedule period: the pre-schedule change period (2013/2014 to 2019/2020 - the 2+1 schedule era) and the post-schedule change period (2020/2021 to 2024/2025 - the 1+1 schedule era). For each schedule period we identified statistical outliers using the interquartile range (IQR) method (i.e. outlier thresholds calculated as the 75^th^ percentile plus 1.5 times the IQR).

### 2.3. Pneumococcal disease susceptibility

To estimate the vulnerability of specific birth cohorts to VT IPD at defined timepoints based on observed vaccine uptake patterns, we used a calculation combining the PCV coverage data with published vaccine effectiveness estimates for PCV13 against IPD. These estimates address susceptibility to VT IPD (i.e. IPD caused by serotypes covered by PCV13), but do not account for changes in risk form non-vaccine serotypes, which may vary over time.

#### 2.3.1. The susceptibility calculation

We classified as susceptible those at risk from contracting VT IPD due to either non-vaccination, partial vaccination (i.e. not completing the full schedule of doses) or the residual risk of disease after full vaccination ^8,20^.

Consequently, for each birth cohort (stratified by quarter) we calculated susceptibility as:

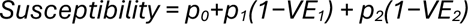

where *p* 0 corresponds to the proportion of children unvaccinated, *p*1 the proportion who received only the primary doses (referred to as the ‘booster gap’ – the difference between the booster vaccine uptake at 24 months in the reference quarter and the primary vaccine uptake at 12 months one year prior) and *p*2 the proportion who received the booster dose - for instances where booster vaccine uptake at 24 months in the reference quarter was greater than primary vaccine uptake at 12 months one year prior, we assumed none of the population had received primary doses only (i.e. *p*1 = 0). VE1 and VE2 denote the estimated vaccine effectiveness against VT IPD after receiving the primary doses and booster dose, respectively.

We assumed that vaccine protection remained constant at the specified effectiveness levels throughout the observation period (up to 24 months post-booster). We acknowledge that our susceptibility calculation does not account for partial catch-up, waning immunity or indirect (herd) protection effects. Nonetheless, it does provide a first-order approximation of the proportion of each cohort that remains vulnerable to IPD based on coverage gaps and estimates of vaccine effectiveness against IPD, thereby allowing comparisons across deprivation quintiles and time periods.

#### 2.3.2. Vaccine eNectiveness assumptions

We selected vaccine e]ectiveness parameters for the susceptibility calculation from the European multi-country study by Savulescu et al. ^21^. We expand below on our assumptions, with the vaccine e]ectiveness values used in the analysis summarised in Table 1.

**Table 1.**
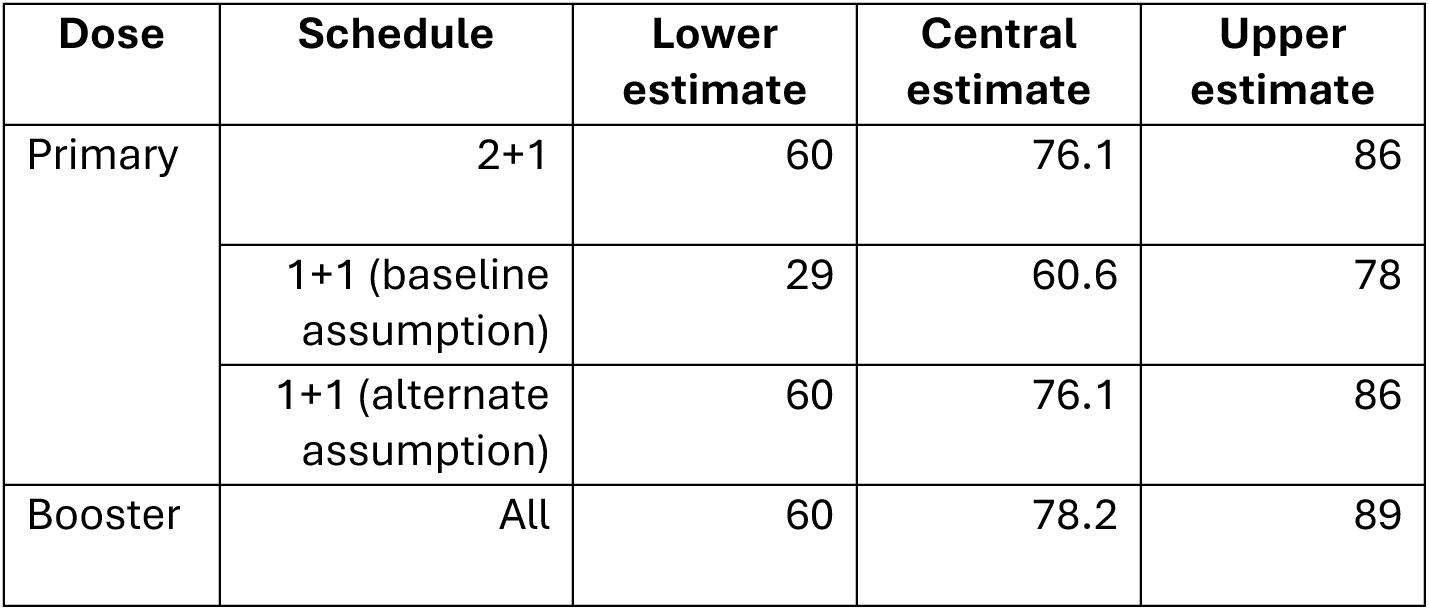
Vaccine e-ectiveness estimates against invasive pneumococcal disease. We list the used lower, central and upper estimates. We sourced the estimates from European based studies reported by Savulescu et al. ^21^.

For the 2+1 schedule, vaccine effectiveness estimates corresponded directly to the published effectiveness data ^21^. Due to limited post-implementation effectiveness data for the 1+1 schedule, vaccine effectiveness estimates required assumptions. For our main analysis, we assumed a vaccine effectiveness of 60.6% after the primary dose; this matched the vaccine effectiveness estimate after one dose in the 2+1 schedule. We refer to that assumption setup as the ‘1+1 (baseline assumption)’ schedule.

To assess the robustness of our findings given this assumption, we considered a ‘1+1 (alternate assumption)’ where the single primary dose administered at 12 weeks in the 1+1 schedule had an effectiveness value matching the vaccine effectiveness post the secondary dose in the 2+1 schedule (76.1%). This assignment reflects a best-case scenario where vaccine administration at 12 weeks, compared to 8 weeks in the 2+1 schedule, generates a more mature immune response functionally equivalent to a second priming dose. We assumed both schedules had an equivalent booster dose effectiveness (78.2%).

To account for uncertainty in VE estimates, we repeated the susceptibility estimation using lower and upper bounds for vaccine effectiveness against IPD reported in the literature. This enabled an exploration of best- and worst-case protection scenarios.

#### 2.3.3. Measured outcomes

For the different dose schedule and vaccine effectiveness scenarios we report: (i) the estimated proportion of susceptible children within a designated strata); (ii) the estimated susceptibility by local authority. In S5 Text we additionally report the estimated cumulative number of susceptible children by quarterly birth cohort.

## 3. Results

### 3.1. Retained dataset characteristics

Our analysis encompassed 149 upper-tier local authorities across England, spanning an 11-year period from 2013/2014 to 2024/2025 and covering 46 quarterly reporting periods. The study population included 7,036,482 eligible children in the 12-month primary dose cohorts (i.e. those children who reached 12 months of age) and 7,235,389 eligible children in the 24-month booster dose cohorts (i.e. those children who reached 24 months of age). The dataset contained 7,003 total observations.

### 3.2. Trends and inequalities in PCV uptake

#### 3.2.1. Temporal trends in PCV uptake

##### National coverage patterns

During the pre-schedule change period (2+1 schedule, 2013-2020), coverage averaged 93.2% at 12-months and 91.2% at 24-months, yielding a booster gap of 2.0%. Following the transition to the 1+1 schedule (2020-2024), while 12-month coverage remained stable at 93.3%, 24-month booster coverage declined to 89.0%. The larger booster gap of 4.4% more than doubled the pre-schedule retention deficit.

##### Local authority patterns

Achievement of the World Health Organization’s 95% coverage target varied substantially between primary and booster doses. For 12-month coverage, 42.1% of quarterly local authority observations (2,846 out of 6,768) met the 95% threshold, with minimal change between schedule periods (43% under the 2+1 schedule versus 40.9% under the 1+1 schedule).

However, booster coverage performance was considerably weaker, with only 17.8% of observations (1,203 out of 6,768) achieving 95% coverage at 24 months. This is a deterioration from 23.3% of observations meeting the 95% target under the 2+1 schedule, falling to just 10.7% under the 1+1 schedule [S4 Text].

##### Deprivation-related inequalities

There were distinct patterns in PCV uptake across deprivation quintiles over the study period. Primary dose coverage at 12 months remained relatively stable across all deprivation quintiles before 2020, with most groups maintaining coverage between 90-95% [Figure 1a]. PCV booster coverage at 24-months was consistently lower than primary dose coverage across all quintiles, with a more pronounced deprivation gradient. Prior to 2020, booster coverage ranged from approximately 90-94% in the least deprived areas to 87-91% in the most deprived quintiles, representing a persistent gap of 2-4 percentage points [Figure 1b].

**Figure 1.**
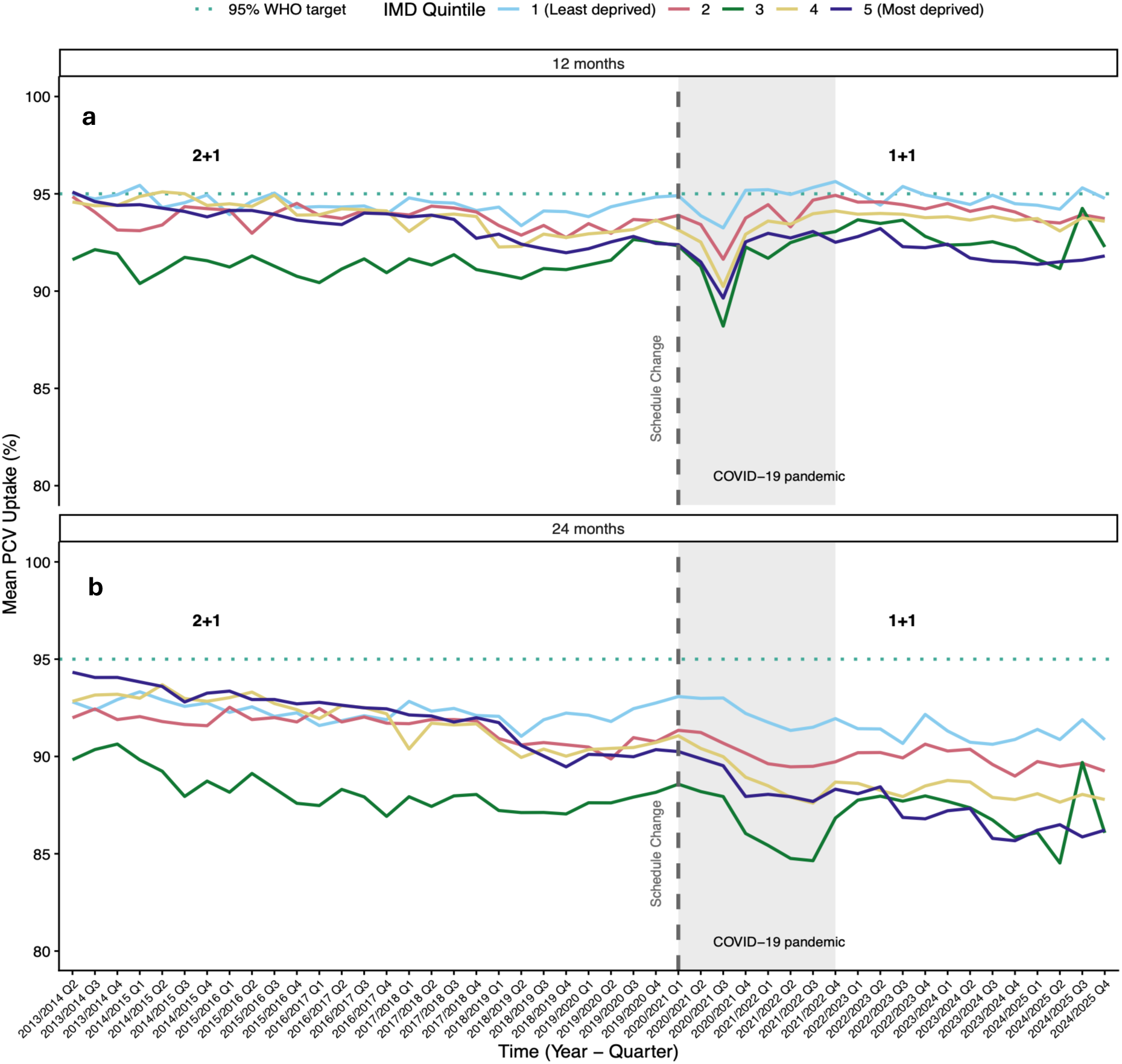
Trends in Pneumococcal Conjugate Vaccine (PCV) uptake at 12 and 24 months by deprivation quintile, 2013–2024. We display for children in England, grouped by deprivation quintile (IMD 1 = least deprived, IMD 5 = most deprived), the: **(a)** average primary dose PCV uptake at 12 months; **(b)** average booster dose uptake at 24 months. The dotted horizontal line marks the WHO-recommended 95% target coverage. The dashed vertical line marks the January 2020 schedule change from a “2+1” to a “1+1” dosing regimen. The shaded grey area highlights the COVID-19 pandemic period. We see a persistent and widening in vaccine uptake over time between the most and least deprived IMD quintiles.

Following the January 2020 schedule change, the COVID-19 pandemic period (2020-2022) caused sharp declines in coverage across all quintiles. The most deprived quintiles (4 and 5) showed larger declines and slower recovery, widening the pre-existing deprivation gap [Figure 1].

By 2023-2024, while 12-month coverage had largely recovered to pre-pandemic levels across most quintiles [Figure 1a], 24-month booster coverage remained depressed, particularly in more deprived areas [Figure 1b]. The deprivation gradient persisted, with quintiles 4 and 5 showing booster coverage 2-3 percentage points below the least deprived quintile - a gap that appears to have widened compared to the pre-2020 period.

As London’s distinct population profile and scale can disproportionately influence national patterns, we generated equivalent results excluding London boroughs. The exclusion of London boroughs from our analysis substantially improved PCV uptake amongst quintile 3 UTLAs; London boroughs comprise 31% of quintile 3 areas but only 7-21% of other quintiles, creating a disproportionate impact on middle-deprivation coverage statistics. The exclusion of London boroughs had a more pronounced impact for booster uptake than primary dose uptake [Figure S4, S4 Text].

#### 3.2.2. Booster retention comparison: Deterioration in the retention of booster doses post the schedule change

In the pre-schedule change period (2+1, 7-year average), Kensington and Chelsea, in London recorded the lowest overall coverage at 70.6% for the minimum of 12-month and 24-month uptake. The local authority with the largest booster gap was Hounslow, London (9.9%). We note that Lancashire recorded a 3.3% higher mean booster uptake by 24-months than mean primary dose coverage at 12-months in the pre-schedule change period. These data indicate potential reporting variations or catch-up vaccination patterns [Figure 2a]. The booster ‘drop-o]’ distribution was heavily concentrated in the 1-2% range, with very few areas experiencing gaps exceeding 8 percentage points [Figure 2c].

**Figure 2.**
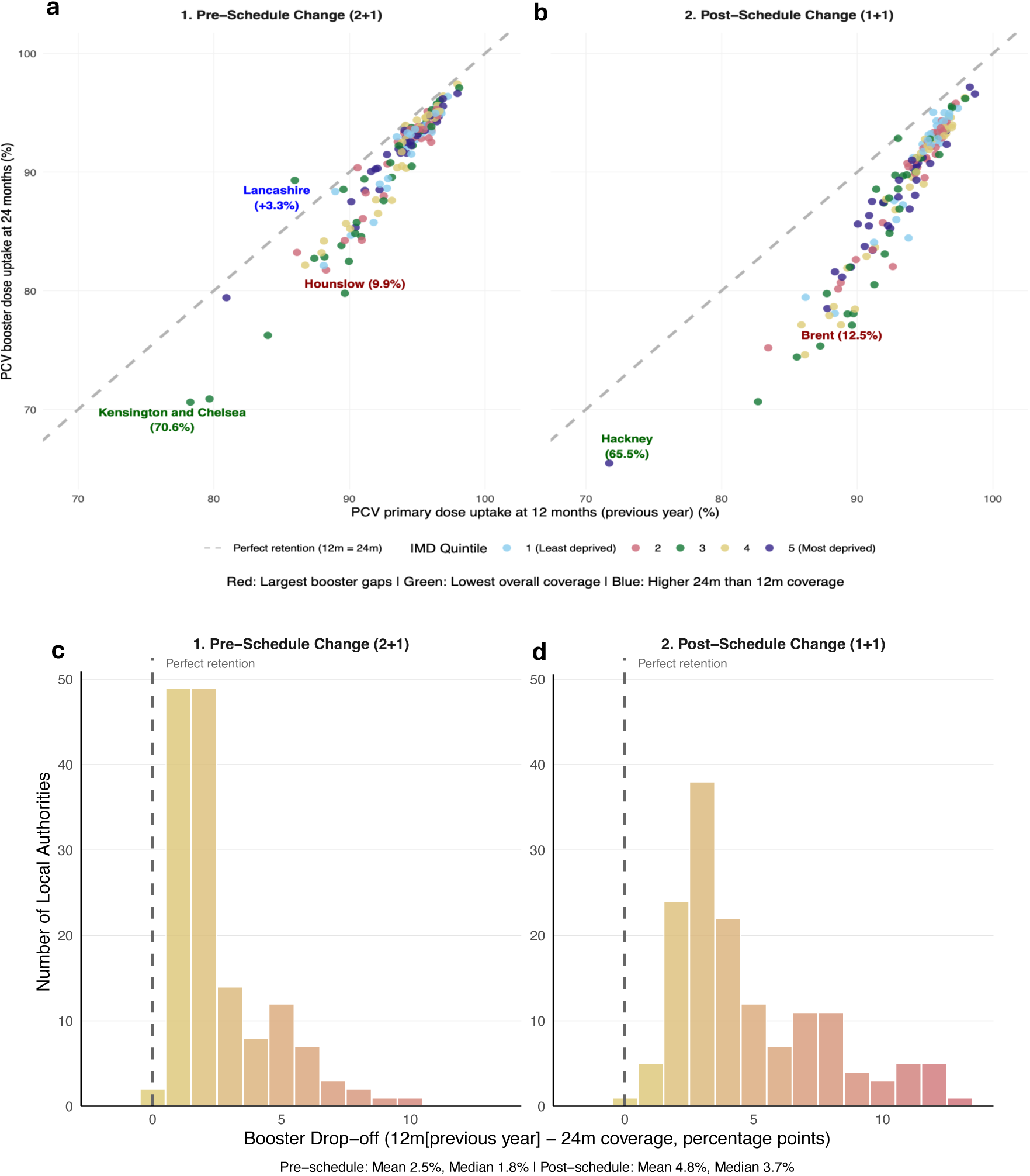
PCV booster retention: Comparison before and after schedule change. (a,b) Each dot represents a local authority. Points are coloured by deprivation quintile. We show average coverage during the: **(a)** 2+1 schedule period (pre-2020, seven-year average); **(b)** 1+1 schedule period (post-2020, five-year average). The dashed grey line shows perfect retention (equal uptake at 12 and 24 months). Dots below the line indicate booster drop-oX. The post-schedule change period has many local authorities falling further below the diagonal, indicating a widespread deterioration in booster completion rates in England. Red labels signify the local authority with the largest booster gap before and after the schedule change (Pre-schedule change: Hounslow, 9.9%; Post-schedule change: Brent, 12.5%). Green labels correspond to the local authorities with the lowest overall coverage. **Blue** labels are local authorities with higher 24-month than 12-month coverage. **(c,d)** Distribution of booster drop-oX during the: **(c)** 2+1 schedule period (pre-2020, seven-year average); **(d)** 1+1 schedule period (post-2020, five-year average). The colour gradient transitions from yellow (lower drop-oX) to red (higher drop-oX rates). The dashed vertical line at zero indicates perfect retention (equal 12-month and 24-month coverage). There is an evident rightward shift in the distribution toward higher drop-oX rates in the post-schedule period.

In the post-schedule change period (1+1, 5-year average) the most concerning booster gaps were observed in Lewisham (11.3%), Greenwich (11.8%), and Brent (12.5%) - all London boroughs showing substantial deterioration in booster retention. Hackney recorded the lowest overall booster coverage at 65.5%, representing further decline in this already disadvantaged area [Figure 2b]. The post-schedule booster ‘drop-o]’ distribution saw the modal bin move to approximately 2.5-3%, whilst becoming more dispersed and extending further into higher drop-o] ranges. The maximum observed gap increased from approximately 9-10% to over 12% [Figure 2d].

These observations confirmed that the aggregate increase in mean booster gaps from 2.32% to 4.79% represented a systematic shift a]ecting many local authorities rather than being driven by a few extreme outliers. Deprivation-related patterns were also evident in both periods. Higher deprivation quintile areas (quintiles 4 and 5) more frequently appeared among the outliers with large booster gaps.

### 3.3. Vaccine Type Invasive Pneumococcal Disease susceptibility

By combining observed vaccination uptake data with published vaccine effectiveness estimates, we estimated the proportion of children who remain susceptible to VT IPD across different time periods, deprivation levels, and geographic areas.

#### 3.3.1. Most deprived populations have an increasing proportion of children susceptible to VT IPD

Susceptibility patterns to VT IPD varied considerably across deprivation quintiles and time periods. Quintile 3 demonstrated the most volatile patterns, with the highest susceptibility peaks exceeding 30% during 2018-2019 and again during 2021-2022. Throughout most of the study period, quintiles 3 and 5 (most deprived) showed the highest susceptibility levels, while quintile 1 (least deprived) generally maintained the lowest susceptibility [Figure 3].

**Figure 3.**
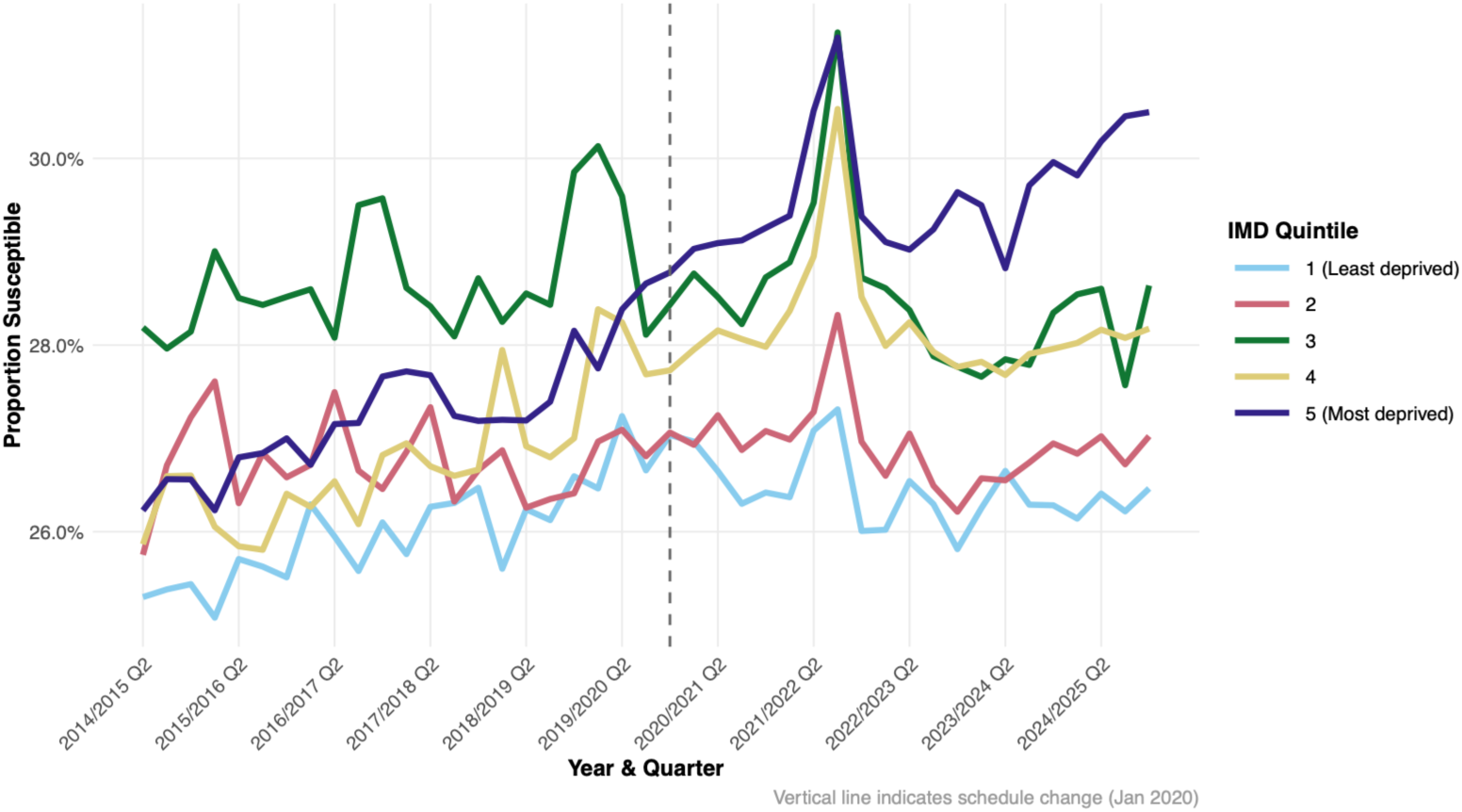
Estimated susceptibility to invasive pneumococcal disease by deprivation quintile: Baseline vaccine e-ectiveness assumption. Lines represent IMD quintiles (1 = least deprived, 5 = most deprived). The vertical dashed line marks the January 2020 schedule change. We observe persistent deprivation gradients and elevated vulnerability in quintiles 3 and 5 throughout the study period.

The 1+1 schedule period demonstrated greater volatility compared to the 2+1 era, with marked fluctuations particularly evident during 2020-2022 (coinciding with the COVID-19 pandemic). The post-2020 period showed increased divergence between quintiles, with the gap between the best and worst performing quintiles widening from approximately 2-3% pre-2020 to 4-5% in later quarters [Figure 3].

#### 3.3.2. Notable geographical variation in estimated VT IPD susceptibility in England

Susceptibility estimates demonstrated marked geographic heterogeneity across England, with local authority-level susceptibility ranging from 22.4% to 47.8% [Figure 4].

**Figure 4.**
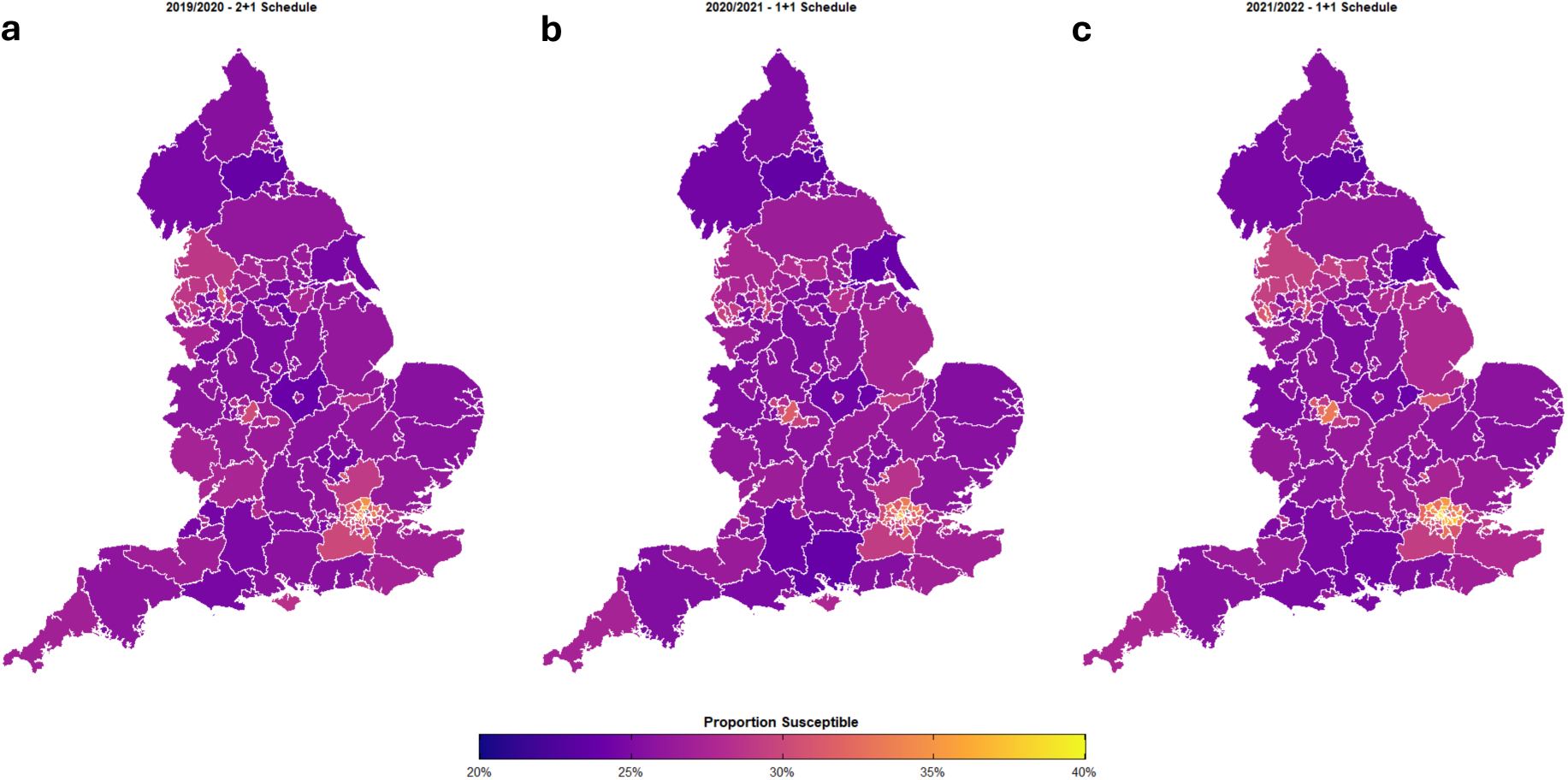
Geographic variation in estimated susceptibility to invasive pneumococcal disease across upper-tier local authorities in England. We show average susceptibility by upper-tier local authority for: (a) 2019/2020 (2+1 schedule); (b) 2020/2021 (transition); (c) 2021/2022 (1+1 schedule). Colour scale: purple (20% susceptibility) to yellow (35% susceptibility). Estimates based on observed coverage and baseline vaccine eXectiveness assumptions (1+1 primary VE = 60.6%). Geographic patterns remain relatively consistent across periods, with most areas showing 20-30% susceptibility. Pockets of higher vulnerability are visible in urban centres, reflecting local variation in vaccination coverage and population characteristics. For the period 2021/2022, we have filled the ’Northamptonshire’ UTLA based on a weighted average from the ’West Northamptonshire’ & ’North Northamptonshire’ data.

During the 2+1 schedule period (2013-2020), most local authorities displayed susceptibility levels between 20-30%, indicating moderate protection levels. There was consistent clustering of well-protected and more vulnerable areas [Figure 4a]. Spatial patterns under the 1+1 schedule (2020-2025) were broadly similar; most local authorities continued to show susceptibility levels in the 20-30% range. Subtle variations in protection levels were evident across regions, but we did not see dramatic geographic polarisation [Figure 4c].

#### 3.3.3. Findings robust to alternate vaccine eNectiveness assumptions

Under the central alternate vaccine effectiveness assumption (1+1 primary dose VE = 76.1%, equivalent to completed 2+1 primary course), the mean population susceptibility within each schedule period and for all local authorities was 27.1% for both the 2+1 schedule period and 1+1 schedule period. That compared to a 0.7 percentage point increase in the mean population susceptibility between schedule periods under the baseline assumption (27.1% for the 2+1 schedule period; 27.8% for the 1+1 schedule period). As observed for the baseline vaccine effectiveness assumption, quintiles 3 and 5 (most deprived) showed the highest susceptibility levels, while quintile 1 (least deprived) generally maintained the lowest susceptibility. The differing VE assumptions also resulted in a similar range in minimum and maximum mean susceptibility values across the local authorities during the 1+1 schedule period (Baseline assumption: 23.2-45.2%; Alternate assumption: 22.9-44.5%) [Figure S6, S6 Text].. Geographic patterns remained consistent across both VE assumptions, with no dramatic changes in regional vulnerability patterns [Figure S7, S6 Text].

Qualitative patterns in susceptibility estimates were maintained when applying either the lower or upper vaccine effectiveness estimates [Table 1]. Quantitatively, comparing susceptibility estimates to when applying the central vaccine effectiveness assumption (where susceptibility estimates across IMD quintiles were between 25-31%), the susceptibility estimates were elevated when applying the lower vaccine effectiveness assumption (between 42-48%; Figure S8, S6 Text) and reduced when applying the higher vaccine effectiveness assumption (between 15-22%; Figure S9, S6 Text).

## 4. Discussion

This study examined trends and inequalities in PCV13 uptake across England following the January 2020 transition from a 2+1 to 1+1 dose regimen. The analysis revealed three critical findings: booster retention deteriorated during the 1+1 schedule; socioeconomic inequalities in vaccine uptake persisted and widened (particularly affecting the most deprived communities); a growing population-level and inequitable susceptibility to VT IPD.

The schedule change in January 2020 coincided with the start of COVID-19 pandemic and it’s associated impacts, including but not limited to the UK’s introduction of PHSMs, widespread health system disruption and burden, and changing vaccine confidence, with associated declines in other paediatric vaccines since 2019. Against this backdrop, the most striking finding from our study was the systematic national deterioration in booster retention following the schedule transition (2.32% for 2+1; 4.79% for 1+1). The scientific literature supporting the schedule change, including work by Ladhani et al. ^5^ and Goldblatt et al. ^22^, demonstrated non-inferiority in controlled trial conditions but could not have adequately anticipated the operational challenges of maintaining booster completion in routine healthcare delivery post COVID-19. Furthermore, there is emerging evidence of “booster hesitancy”, even among those who accepted initial vaccination. The terminology itself may be problematic - the gap between “fully vaccinated” status (after primary doses) and the need for a “booster” creates cognitive dissonance. Lin et al. ^23^ found that vaccinated-but-not-boosted individuals expressed confusion about the need for additional doses and discontentment that this requirement was not communicated from the start. Additionally, because the PCV booster is administered alongside the first dose of the Measles, Mumps, and Rubella (MMR) vaccine, its receipt may be indirectly influenced by vaccine hesitancy stemming from the post-Andrew Wakefield MMR scandal ^24,25^.

Second, although the deprivation gradient was consistently present across both schedule periods, the gap between the least and most deprived quintiles expanded from approximately 2-3% pre-2020 to 4-6% in later quarters. These data corroborate previous research demonstrating widening socioeconomic disparities across multiple paediatric vaccines during the post COVID-19 pandemic period ^8^. Our observation that quintile 3 areas often performed worse than the most deprived (quintile 5) requires careful interpretation. This is partially an effect of London local authorities (many in quintile 3 [S3 Text]) having the widest internal heterogeneity in deprivation levels, including some of the most deprived neighbourhoods in England.

Third, susceptibility estimates suggest that the burden of children vulnerable to VT IPD by 2024/25 disproportionately concentrated in disadvantaged areas. The pneumococcal vaccination programme provides population-level protection through both direct immunity in vaccinated individuals and indirect (herd) immunity that particularly benefits vulnerable populations including the elderly. This dual protection mechanism means that declining vaccine uptake in children can have cascading effects on community-wide disease prevention. Higher valency PCVs with reduced potential for carriage control as a consequence of lower immunogenicity may compound this ^26^. Areas with higher levels of socioeconomic deprivation may experience more rapid erosion of herd immunity due to lower vaccination coverage, potentially widening health inequalities that extend beyond the directly affected paediatric population to encompass older vulnerable groups who rely on community protection.

These findings must, however, be interpreted in the context of study assumptions and limitations. The first limitation relates to the use of UTLA-level deprivation measures and vaccination uptake statistics. UTLAs encompass diverse neighbourhoods ranging from affluent to severely deprived. Area-based deprivation indices can therefore fail to capture significant proportions of deprived individuals ^27^, exemplified in the London boroughs. This spatial aggregation may obscure important local patterns of vaccination uptake in pockets of deprivation within otherwise affluent areas, or conversely areas of relative advantage within deprived authorities. Ideally, we would conduct the analysis at GP practice level, which would better capture local deprivation patterns and their relationship with vaccination uptake. However, GP-level vaccination data were not available before 2018 for the 2+1 regimen period.

Second, the susceptibility calculation relied on vaccine effectiveness estimates derived from European studies of the 2+1 schedule. Additionally, the assumptions about 1+1 effectiveness have not yet been fully validated in post-implementation surveillance. Given that uncertainty, our sensitivity analysis can offer insight on a plausible range of estimates for susceptibility. Of interest for further study are sensitivity analyses using distributional estimates of vaccine effectiveness, which may be performed using probabilistic modelling. More complex susceptibility models may be devised that consider both direct protection from receiving the vaccine and indirect protection via herd immunity. Further processes could be added to account for possible waning immunity; we assumed constant protection from vaccination through time, whereas relaxing that assumption would result in a rise in susceptibility amongst the population.

The third limitation is the susceptibility calculation assumes children remain in their assigned vaccination status without accounting for potential catch-up vaccination. An analysis of timeliness of childhood vaccination in England using primary care electronic health records for children born between 2006 and 2014 showed that by five years of age there was a couple of percentage points increase in PCV primary dose uptake and booster dose uptake ^28^. Although the COVER data not reporting PCV uptake percentages at later ages affects the precision of our booster retention calculations, the observed patterns remain valid for assessing population-level trends in vaccination completion.

Finally, the observed deprivation-uptake associations could be influenced by confounding factors at the area level, including ethnicity, sex, gender, long-term health conditions and religion. Though the ecological study design provides implementation advantages due to its simplicity, it does preclude causal inference about individual-level relationships. The schedule change coinciding with the onset of the COVID-19 pandemic also presents challenges in disentangling the effect of the new schedule from pandemic-related disruptions on PCV uptake.

## 5. Conclusion

This study demonstrates that PCV booster retention has deteriorated substantially in England since the schedule change, which coincided with the start of the COVID-19 pandemic. This deterioration disproportionately affects children in more deprived areas, risking avoidable disease burden concentrated in the most disadvantaged communities and widening health inequalities. Immunisation system strengthening, targeted, equity-focused interventions and enhanced call-recall systems for post-infant vaccine delivery are needed address the identified coverage gaps (see Box 1 for our policy and practice recommendations). Before altering vaccine schedules in other European countries, inequalities in vaccine uptake and booster retention, must be factored into predictions of population impact, alongside measures of immunogenicity, VE estimates and cost-effectiveness. Otherwise, the full protective potential of the 1+1 schedule may not be realised.

##### Box 1: Policy and practice recommendations

The deterioration in booster retention is evident across multiple measures and geographic areas. The deprivation gradient is consistent with established patterns in childhood vaccination and the susceptibility estimates provide a reasonable first-order approximation of population protection gaps based on available evidence.

Several evidence-based recommendations emerge based on these findings.

###### Enhanced follow-up systems

Call-recall systems that were sufficient when children had higher protection from two priming doses may be inadequate when a single primary dose provides more limited early protection. We suggest investment in robust booster dose call-recall systems, particularly in areas with demonstrated large booster gaps. This should include automated reminders, integration across GP and health visitor services and targeted outreach in high-risk areas.

###### Equity-focused interventions

Implement targeted interventions in deprived areas, including mobile vaccination units, community-based clinics, and culturally appropriate reminder systems. Standard approaches appear insufficient to address the persistent deprivation gradient.

###### Geographically targeted interventions

Focus resources on areas with consistently poor performance, particularly London boroughs and other urban areas with large booster gaps.

###### Implementation research

Conduct rigorous evaluation of the 1+1 schedule’s real-world effectiveness, including linkage of coverage data with disease outcomes. Existing inequalities in vaccine uptake and booster retention should be assessed prior to any schedule changes. This should inform evidence-based decisions about potential programme modifications.

###### Real-time monitoring

Establish routine monitoring of booster gaps as a key performance indicator for the vaccination programme. This should include regular reporting by deprivation quintile and geographic area to enable early identification of emerging problems.

## Supporting information

Tracked changes document.

## Data Availability

All data utilised in this study are publicly available. We state relevant references and data repositories stated within the main manuscript.
Data and code associated with the study are available in the following repository: https://github.com/aaliswalker/DASC500-Dissertation---Inequalities-in-Childhood-Pneumococcal-Vaccine-Uptake-in-England
Archived code for this version of the manuscript: https://doi.org/10.5281/zenodo.17854535

https://github.com/aaliswalker/DASC500-Dissertation---Inequalities-in-Childhood-Pneumococcal-Vaccine-Uptake-in-England

https://doi.org/10.5281/zenodo.18420636

## Author contributions

**Praise Ilechukwu:** Data curation, Formal analysis, Methodology, Software, Validation, Visualisation, Writing - Original Draft, Writing - Review & Editing.

**Daniel Hungerford:** Conceptualisation, Methodology, Supervision, Validation, Visualisation, Writing - Original Draft, Writing - Review & Editing.

**Neil French:** Conceptualisation, Methodology, Writing - Review & Editing.

**Edward M. Hill:** Conceptualisation, Methodology, Supervision, Validation, Visualisation, Writing - Original Draft, Writing - Review & Editing.

## Data and code availability statement

All data utilised in this study are publicly available. We state relevant references and data repositories stated within the main manuscript.

Data and code associated with the study are available in the following repository: https://github.com/aaliswalker/DASC500-Dissertation---Inequalities-in-Childhood-Pneumococcal-Vaccine-Uptake-in-England

Archived code: https://doi.org/10.5281/zenodo.18420636

## Supplementary Material

### S1 Text. COVER data preparation

We downloaded all pneumococcal vaccine uptake data from the COVER programme for the years 2013 to 2024, spanning a total of 52 quarterly files. These files varied in structure, format (.xlsx, .ods), and naming conventions. To ensure consistency, we renamed all files using a uniform format: 20XX_QX for pre-2020 files and 20XX QX for post-2020 files.

We imported all quarterly PCV uptake files into R and combined them into a single dataset for analysis. Due to inconsistencies across years—including changes in sheet names, column headers, and data formatting—we wrote custom cleaning scripts to extract only the relevant PCV tables (i.e., 12-month and 24-month PCV coverage and population denominators). We filtered out unrelated tables (e.g. MMR, Hib/MenC), and harmonised all extracted columns using a consistent naming convention (e.g., PCV_12m, PCV_24m, ONS_Code). [Table S1]

To prevent data type mismatches, we initially read all columns as character vectors, before converting them to appropriate types for analysis (e.g., numeric for coverage percentages and denominators, factor for categorical variables). To ensure that we handled missing values consistently, we recoded any value starting with ’N’ (e.g., “N.A.”) or containing extraneous symbols (such as those beginning with ’[’ e.g [z]) as NA.

**Table S1.**
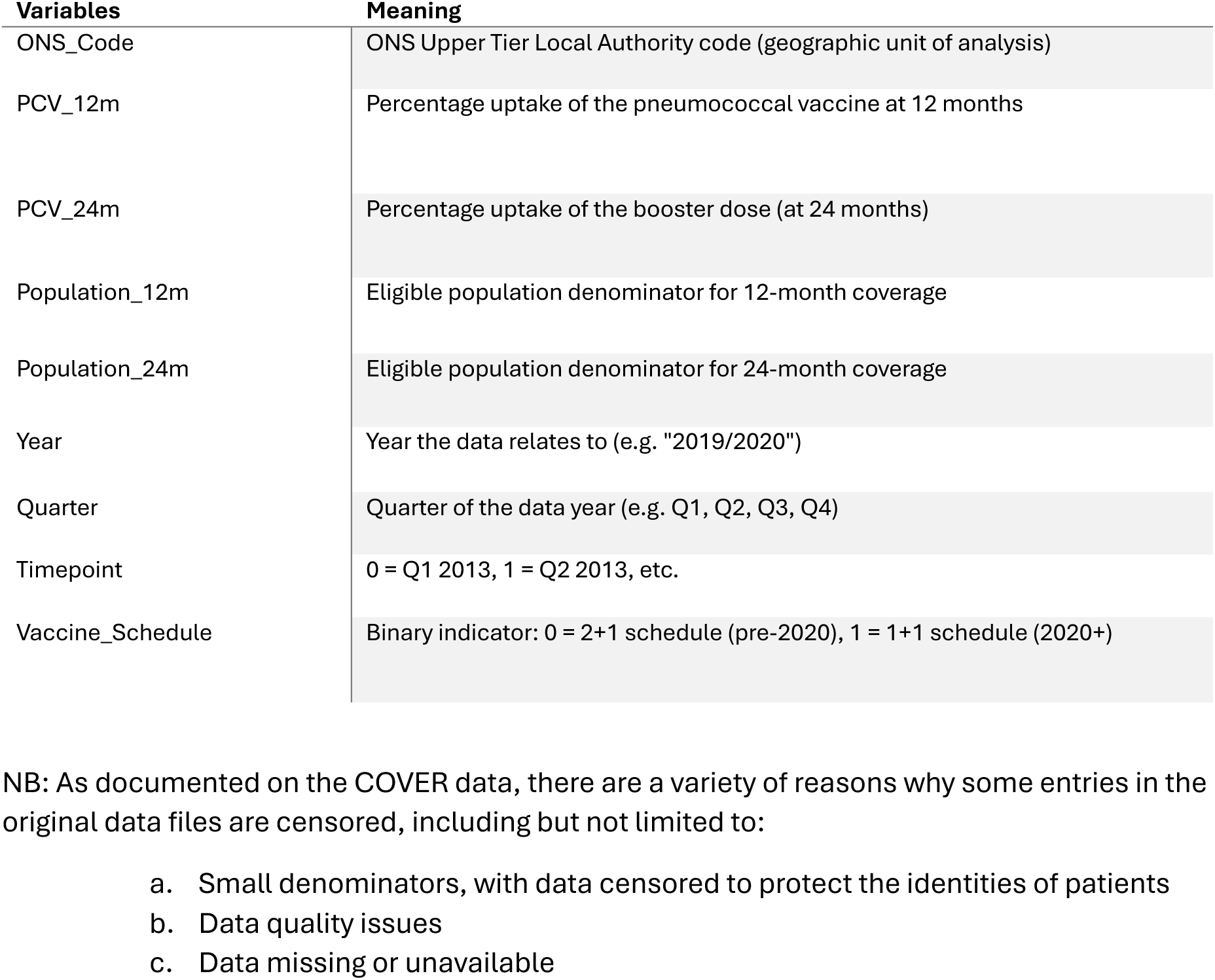
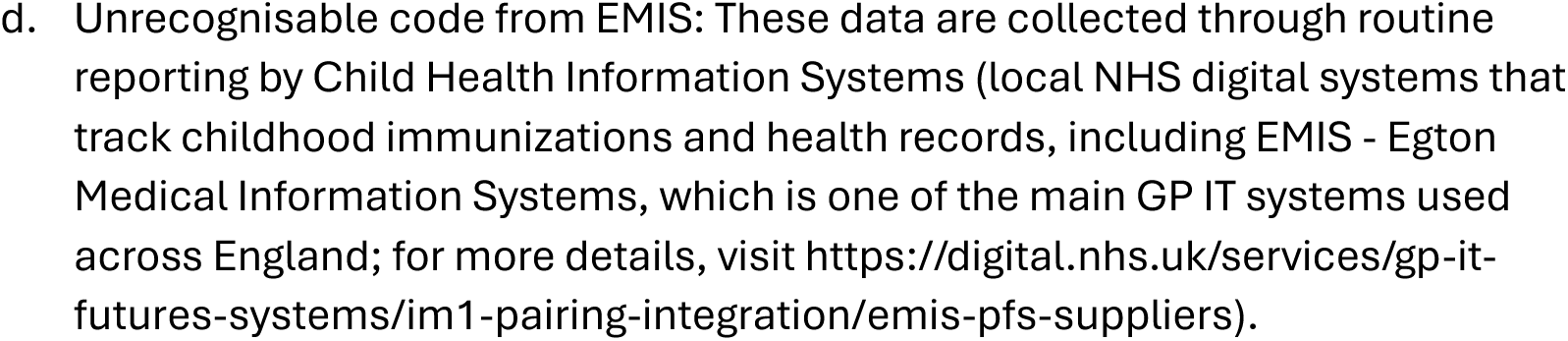
Variables used for data preparation.

### S2 Text. COVER data cleaning

Cross-referencing quarterly COVER files against the 2019 Index of Multiple Deprivation UTLA list identified two authorities that did not appear in all years: Bournemouth, Christchurch and Poole (E06000058) and Northamptonshire (E10000021).

- **Bournemouth, Christchurch and Poole (E06000058):** This authority code first appeared in the COVER dataset from 2019/2020 onwards. It was absent from COVER data for 2013/2014 through 2018/2019, where instead there were two separate authorities: Bournemouth (E06000028) and Poole (E06000029).
- **Northamptonshire (E10000021):** This authority code appeared in COVER data from 2013/2014 through 2020/2021. It was absent from the dataset from 2021/2022 onwards. During this same period, two new authority codes appeared: West Northamptonshire (E06000061) and North Northamptonshire (E06000062).

We also identified two authorities whose ONS code changed during the study period: Buckinghamshire and Dorset.

- **Buckinghamshire:** We verified ONS code consistency across all years and standardised records to the current code (E06000060).
- **Dorset:** Early COVER files used code E10000009, while later files (2020 onwards) used code E06000059 for the same geographic area. We standardised all records to use E06000059 to ensure temporal consistency.

These temporal variations resulted in 149 distinct UTLA codes appearing across the study period, though not all 149 appeared in every quarterly file. The final analytical dataset comprised 6,816 observations with no records excluded due to missing or invalid geographic identifiers.

### S3 Text. UTLA stratification by IMD quintile

We merged our cleaned vaccine uptake dataset with the IMD dataset using the ONS code as a key. We retained only matched records. We then split IMD scores into quintiles using the ntile() function, with quintile 1 corresponding to the least deprived and quintile 5 to the most deprived local authorities.

We list below the UTLA’s that were classified under each IMD quintile. These classifications used UTLA-level Index of Multiple Deprivation (IMD) summary data from the English Indices of Deprivation 2019 ^15^. For each IMD quintile we list the UTLA’s alphabetically.

#### IMD quintile 1 (Least Deprived)

Bath and North East Somerset, Bracknell Forest, Bromley, Buckinghamshire, Cambridgeshire, Central Bedfordshire, Cheshire East, Dorset, East Riding of Yorkshire, Gloucestershire, Hampshire, Harrow, Hertfordshire, Kingston upon Thames, Leicestershire, Merton, North Yorkshire, Oxfordshire, Richmond upon Thames, Rutland, South Gloucestershire, Surrey, Sutton, Warwickshire, West Berkshire, West Sussex, Wiltshire, Windsor and Maidenhead, Wokingham, York.

#### IMD quintile 2

Barnet, Bedford, Bexley, (Bournemouth, Christchurch and Poole*), Camden, Cheshire West and Chester, Derbyshire, Devon, Dorset, East Sussex, Essex, Havering, County of Herefordshire, Hillingdon, Kent, Milton Keynes, North Somerset, Northamptonshire^, Nottinghamshire, Reading, Redbridge, Shropshire, Solihull, Somerset, Sta]ordshire, Su]olk, Swindon, Tra]ord, Wandsworth, Warrington, Worcestershire.

#### IMD quintile 3

Brent, Brighton and Hove, Bury, Cornwall, Coventry, Croydon, Cumbria, Dudley, Ealing, Greenwich, Hammersmith and Fulham, Hounslow, Isle of Wight, Kensington and Chelsea, Kirklees, Lambeth, Lancashire, Lincolnshire, Medway, Norfolk, North Lincolnshire, North Tyneside, Northumberland, Slough, Southend-on-Sea, Stockport, Telford and Wrekin, Thurrock, Waltham Forest, Westminster.

#### IMD quintile 4

Barnsley, City of Bristol, Calderdale, County Durham, Darlington, Derby, Enfield, Gateshead, Haringey, Islington, Leeds, Lewisham, Luton, Newcastle upon Tyne, Newham, Peterborough, Plymouth, Portsmouth, Redcar and Cleveland, Rotherham, Sefton, Sheffield, Southampton, Southwark, Stockton-on-Tees, Torbay, Tower Hamlets, Wakefield, Wigan, Wirral.

#### IMD quintile 5 (Most Deprived)

Barking and Dagenham, Birmingham, Blackburn with Darwen, Blackpool, Bolton, Bradford, Doncaster, Hackney, Halton, Hartlepool, City of Kingston upon Hull, Knowsley, Leicester, Liverpool, Manchester, Middlesbrough, North East Lincolnshire, Nottingham, Oldham, Rochdale, Salford, Sandwell, South Tyneside, St. Helens, Stoke-on-Trent, Sunderland, Tameside, Walsall, Wolverhampton.

*For analysis of data from 2013 to 2019, we applied the IMD quintile assignment for ‘Bournemouth, Christchurch and Poole’ to ‘Bournemouth’ and ‘Poole’ (i.e. IMD quintile 2)

^For analysis of data from 2021 to 2025, we applied the IMD quintile assignment for ‘Northamptonshire to ‘North Northamptonshire’ and ‘West Northamptonshire’ (i.e. IMD quintile 2).

**Figure S1.**
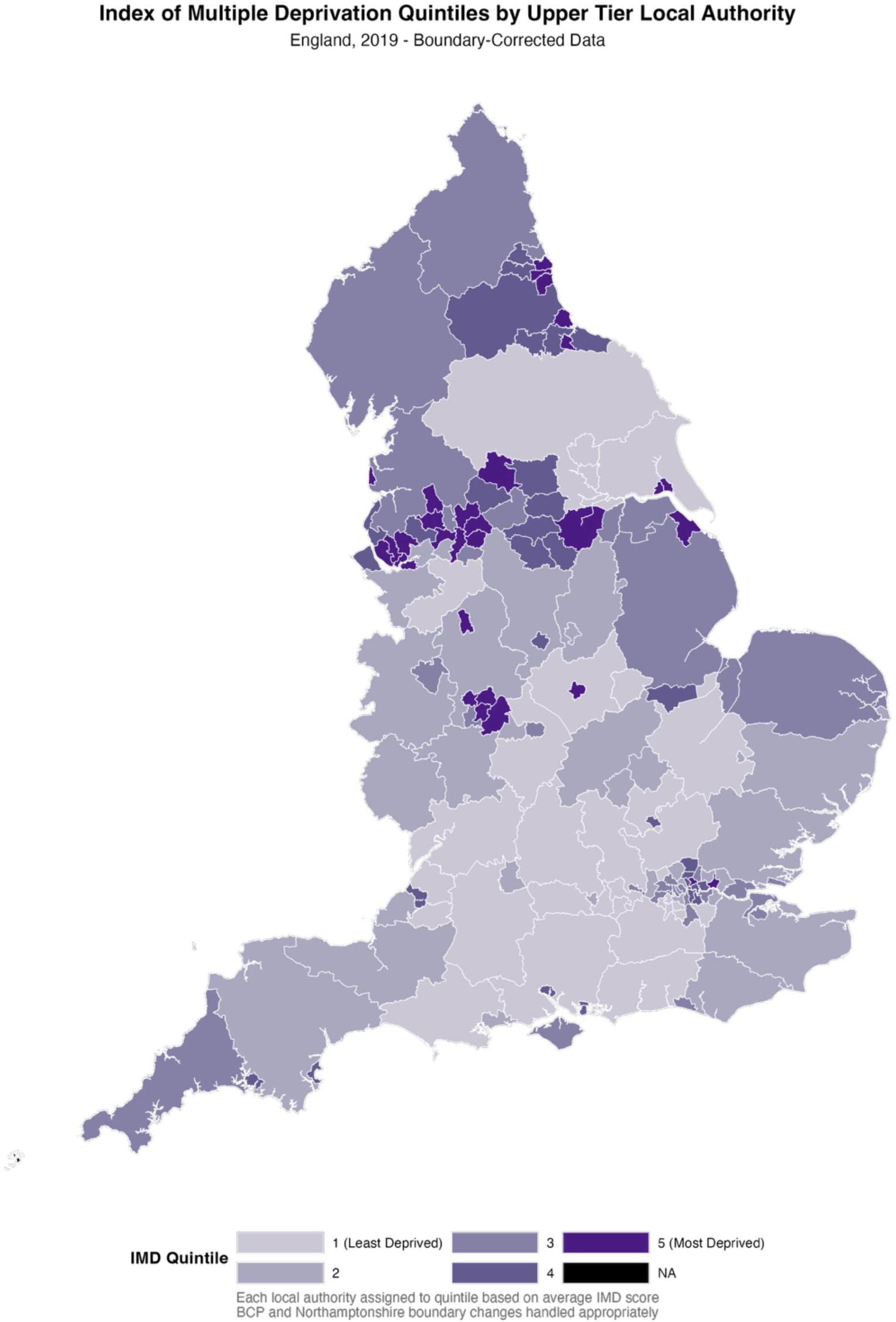
Assignment of UTLAs in England to IMD quintiles. These classifications used UTLA-level Index of Multiple Deprivation (IMD) summary data from the English Indices of Deprivation 2019 15. We assigned each UTLA to an IMD quintile based on the average IMD score for each UTLA. Shading denotes the IMD quintile associated with that UTLA, with the lightest shading representing the least deprived UTLAs (quintile 1) and the darkest shading representing the most deprived UTLAs (quintile 5). We assigned as NA the UTLAs not retained for our study (Isle of Scilly, The City of London).

### S4 Text. Additional PCV uptake figures

**Figure S2.**
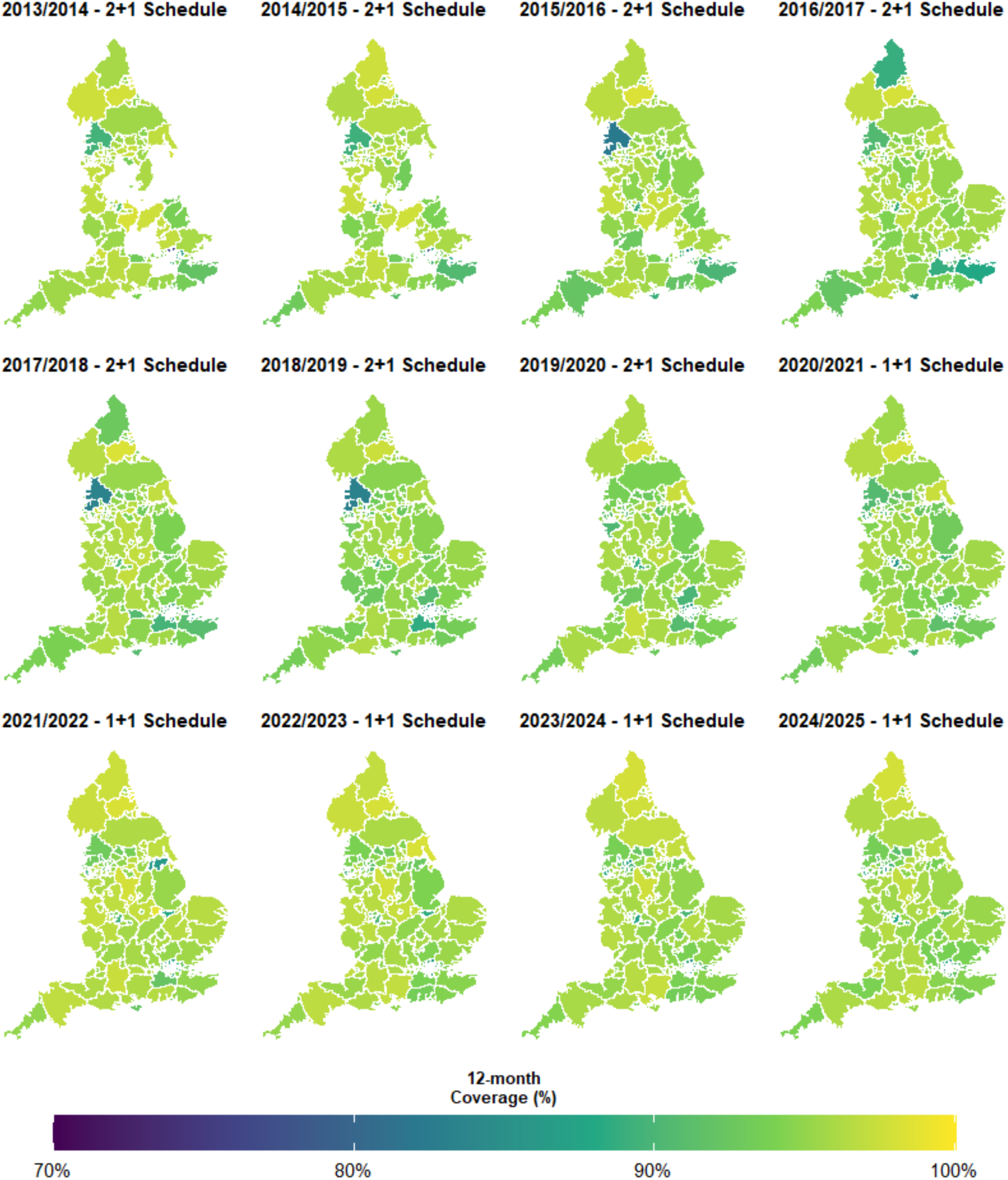
Geographic variation in PCV primary coverage (12 months) across local authorities in England, 2013/2014 to 2024/2025. The colour scale ranges from dark blue/purple (70% coverage) to bright green/yellow (100% coverage), so brighter areas indicate higher vaccination rates while darker areas show lower uptake. The maps are arranged chronologically from left to right, top to bottom, spanning both the 2+1 schedule period (2013-2020), and the 1+1 schedule period (2021-2025).

**Figure S3.**
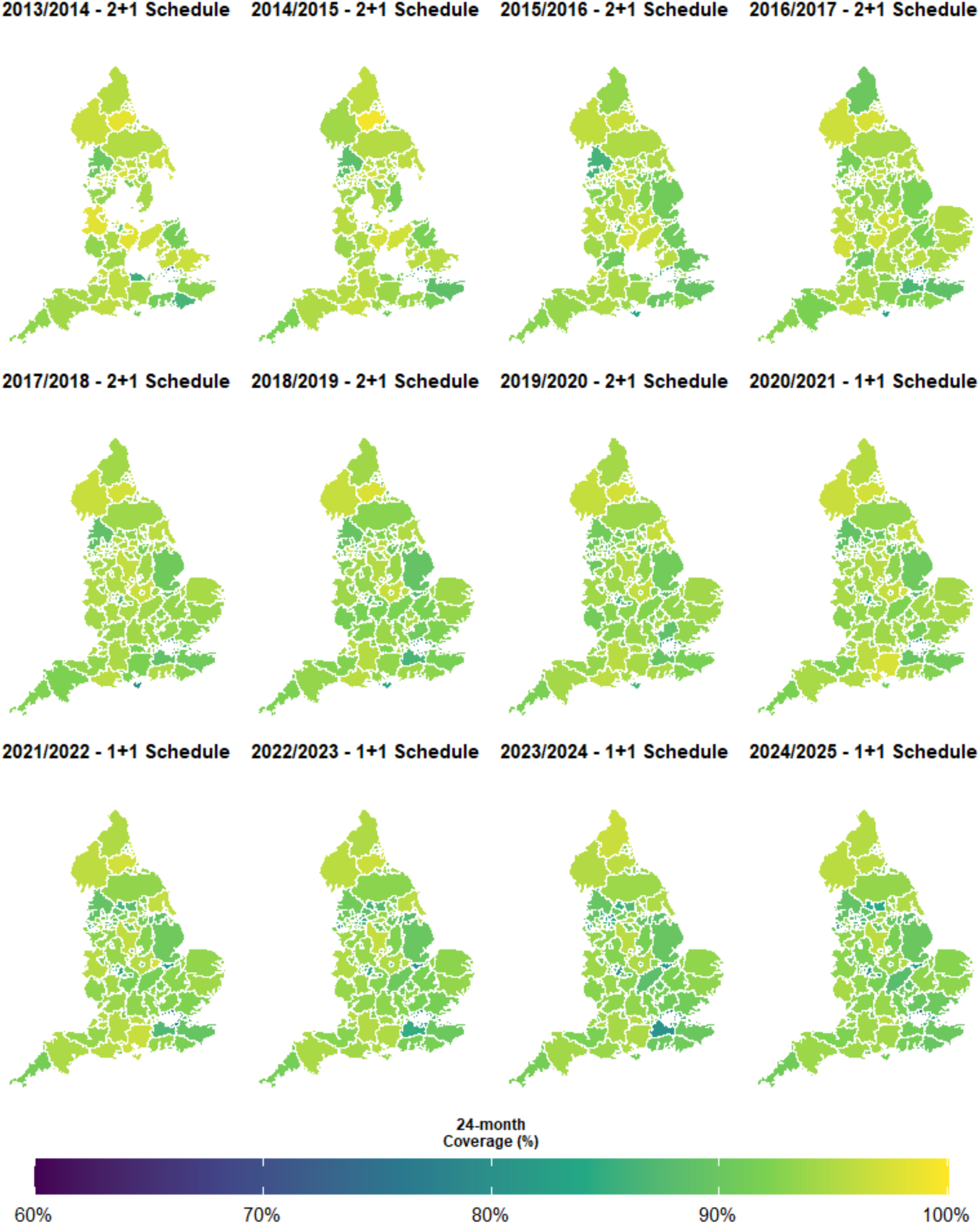
Geographic variation in PCV booster coverage (24 months) across local authorities in England, 2013/2014 to 2024/2025. The colour scale ranges from dark blue/purple (60% coverage) to bright yellow (100% coverage), where brighter green/yellow areas indicate higher booster uptake and darker blue areas show lower coverage. The maps are arranged chronologically from left to right, top to bottom, spanning both the 2+1 schedule period (2013-2020), and the 1+1 schedule period (2021-2025).

**Figure S4.**
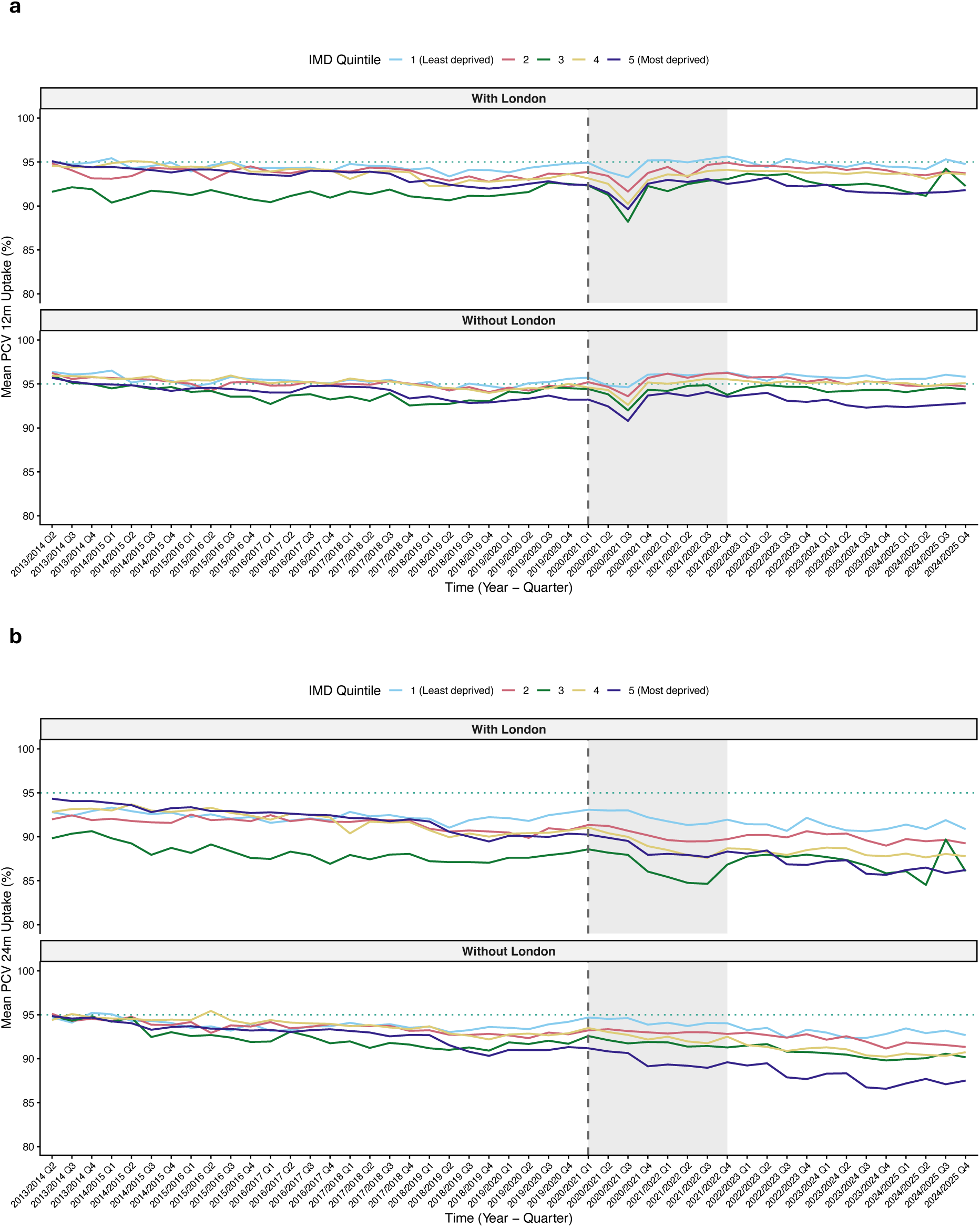
Impact of London on PCV Coverage by deprivation quintile. Comparison of uptake trends for: **(a)** primary doses at 12 months; **(b)** booster dose at 24 months. In each panel, the upper plot includes 32 London boroughs (i.e. all London boroughs except the City of London), whereas the lower panel does not include any London boroughs. Lines represent IMD quintiles (1 = least deprived, 5 = most deprived). Dashed vertical line: schedule change (January 2020); shaded area: COVID-19 period. The impact of London exclusion is more pronounced for booster coverage than primary doses.

### S5 Text. Cumulative VT IPD susceptibility estimates indicate widening socioeconomic inequalities

To better understand the growing burden of disease risk over time, we calculated the cumulative number of susceptible children by birth cohort (quarterly). This analysis demonstrates how even small reductions in uptake can lead to a steady accumulation of susceptible individuals, which may have implications for herd protection and future disease resurgence if left unaddressed [Figure S5a]. Susceptibility patterns revealed persistent and widening inequalities by deprivation level. Although overall susceptibility levels appeared similar before and after the schedule change, the more deprived areas (quintiles 3 and 5) experienced disproportionately higher vulnerability throughout the study period [Figure S5b].

**Figure S5.**
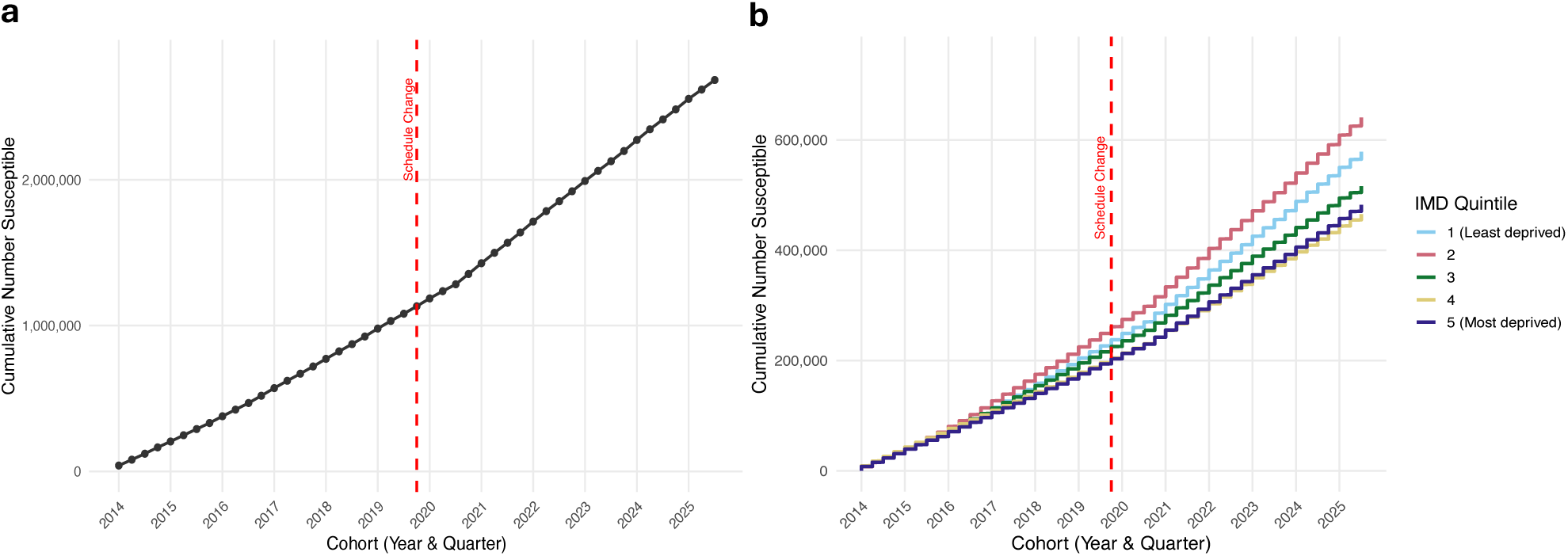
Cumulative number of susceptible children by quarter. We calculated susceptibility on observed uptake and literature-based vaccine eXectiveness. **(a)** Each point represents the total number of children estimated to remain susceptible to invasive pneumococcal disease (IPD) after vaccination, cumulatively summed from 2013/2014 Q2 to 2024/2025 Q3. **(b)** Stratification by deprivation quintiles. (1 = least deprived, 5 = most deprived). The x-axis tracks cohort entry by year and quarter.

### S6 Text. Findings robust to alternate vaccine eQectiveness assumptions

When applying the alternate central vaccine effectiveness assumption in our susceptibility calculation, throughout most of the study period, we found quintiles 3 and 5 (most deprived) had the highest susceptibility levels, while quintile 1 (least deprived) generally maintained the lowest susceptibility [Figure S6]. Geographic patterns remained consistent across both VE assumptions, with similar spatial distributions of susceptibility levels and no dramatic changes in regional vulnerability patterns [Figure S7].

When applying either the lower or upper vaccine effectiveness estimates, rather than the central vaccine effectiveness estimates [Table 1], we found qualitative patterns were maintained [Figures S8 & S9]. Quantitatively, comparing susceptibility estimates to when applying the central vaccine effectiveness assumption (where susceptibility estimates across IMD quintiles were between 25-31%), the susceptibility estimates were elevated when applying the lower vaccine effectiveness assumption (between 42-48%; Figure S8) and reduced when applying the higher vaccine effectiveness assumption (between 15-22%; Figure S9).

**Figure S6.**
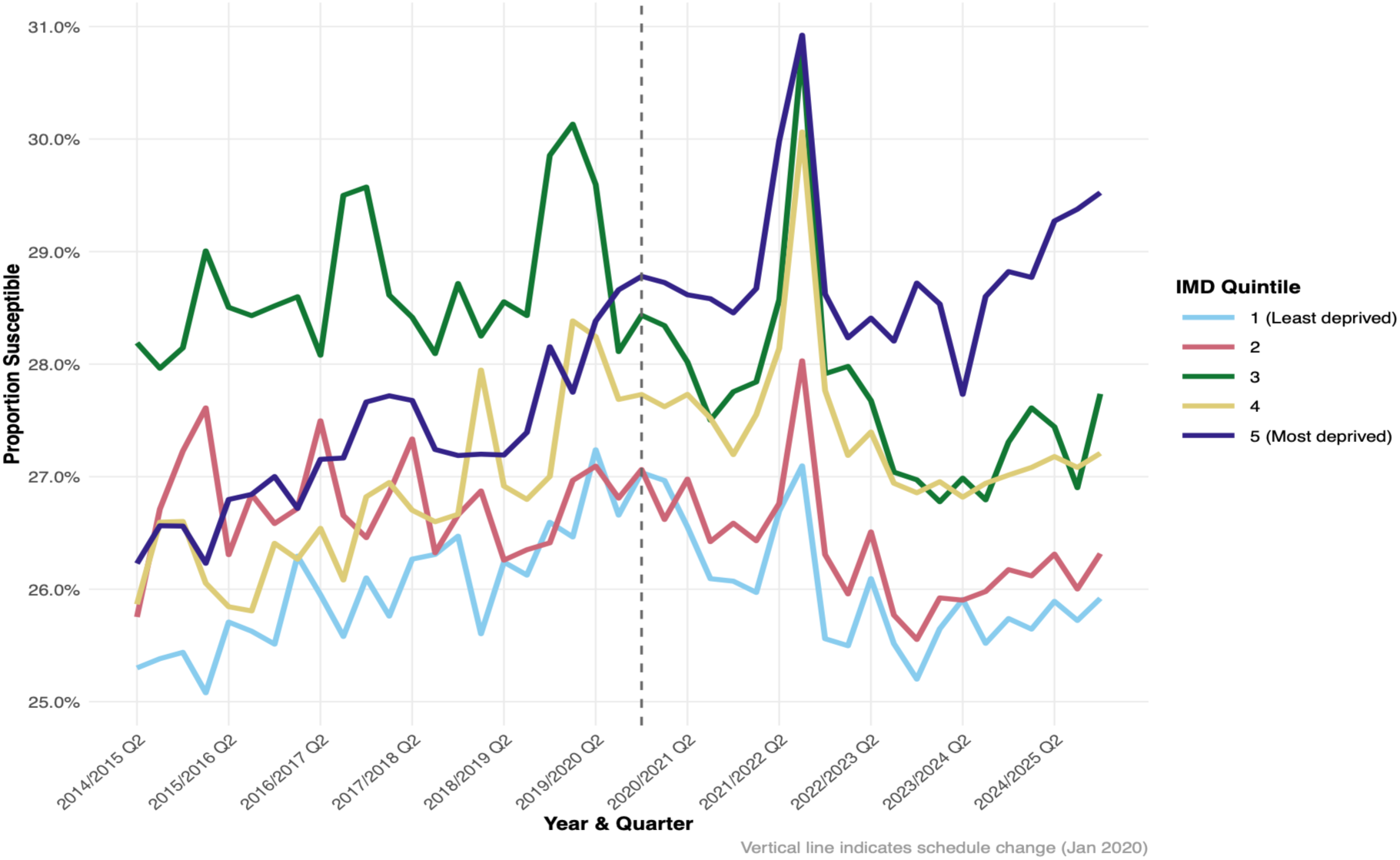
Estimated susceptibility to invasive pneumococcal disease by deprivation quintile: Alternate central vaccine eNectiveness assumption. (1+1 primary dose VE = 76.1%). Lines represent IMD quintiles (1 = least deprived, 5 = most deprived). The vertical dashed line marks the January 2020 schedule change. We observe persistent deprivation gradients and elevated vulnerability in quintiles 3 and 5 throughout the study period.

**Figure S7.**
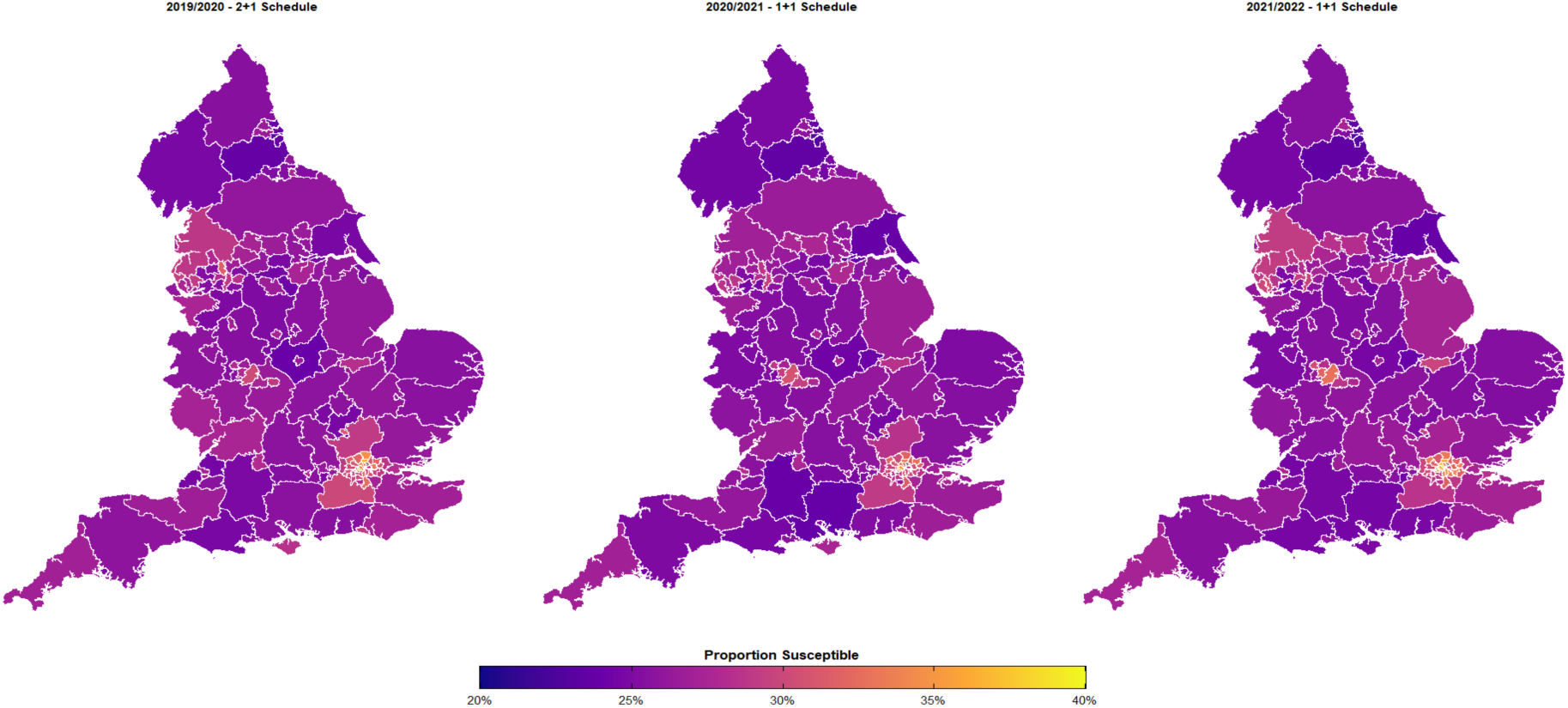
Geographic variation in IPD susceptibility: Alternate central vaccine eNectiveness assumption. We show average susceptibility by upper-tier local authority for 2019/2020 (2+1 schedule), 2020/2021 (transition), and 2021/2022 (1+1 schedule). Colour scale: purple (20% susceptibility) to yellow (35% susceptibility). Estimates based on alternate vaccine eXectiveness assumption (1+1 primary VE = 76.1%, matching completed 2+1 primary course with booster 78.2%). Most areas show 20-30% susceptibility, with persistent pockets of higher vulnerability in urban centres.

**Figure S8.**
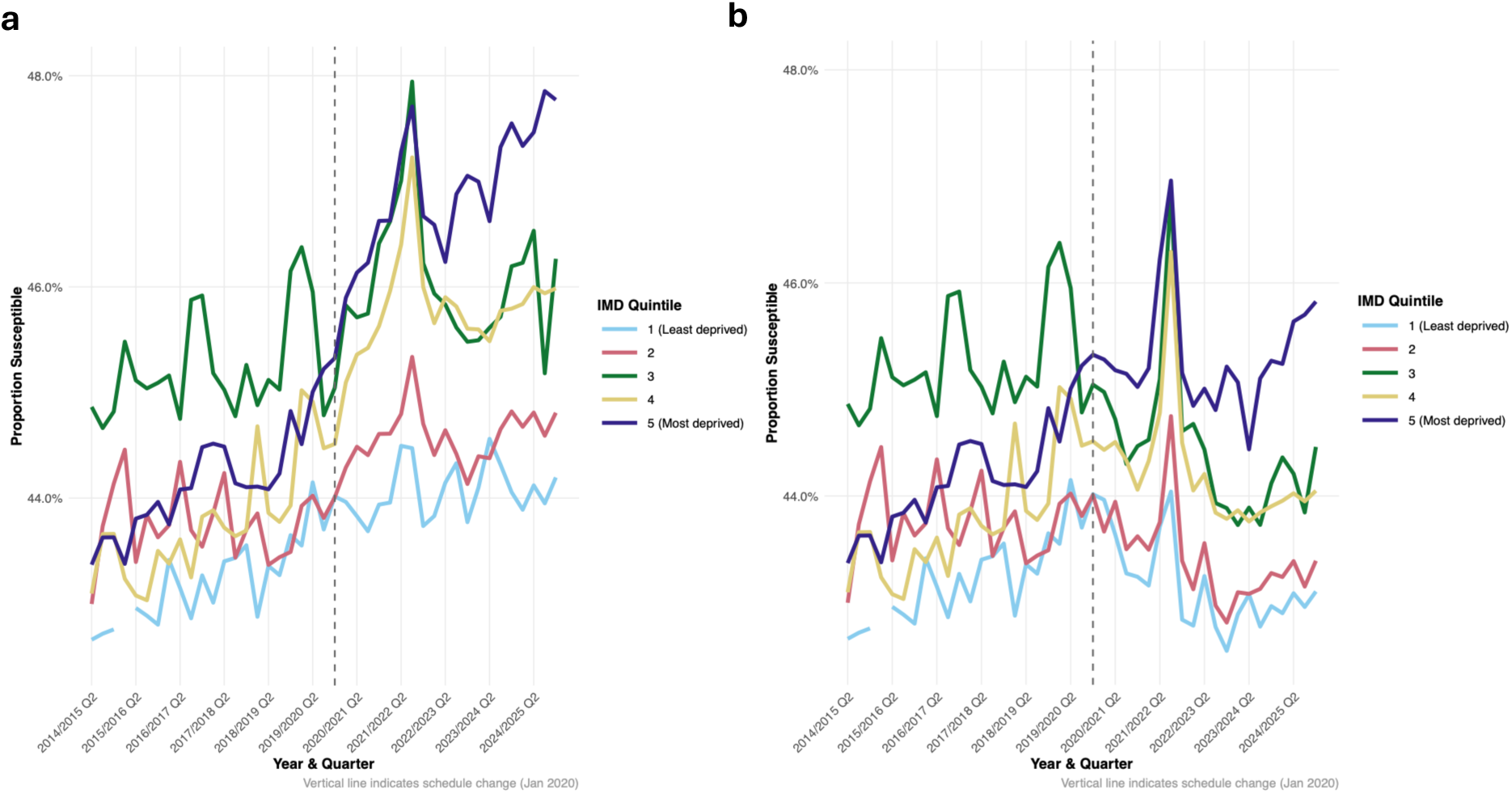
Estimated susceptibility to invasive pneumococcal disease by deprivation quintile: Baseline vs Alternate lower vaccine eNectiveness assumptions. **(a)** Baseline lower vaccine eXectiveness assumption. (b) Alternate lower vaccine eXectiveness assumption. Both scenarios show qualitative similar patterns to the estimates for the central vaccine eXectiveness assumptions. Quantitatively, for both the baseline and alternate scenarios the susceptibility estimates are elevated when applying the lower vaccine eXectiveness assumption compared to when applying the central vaccine eXectiveness assumption.

**Figure S9.**
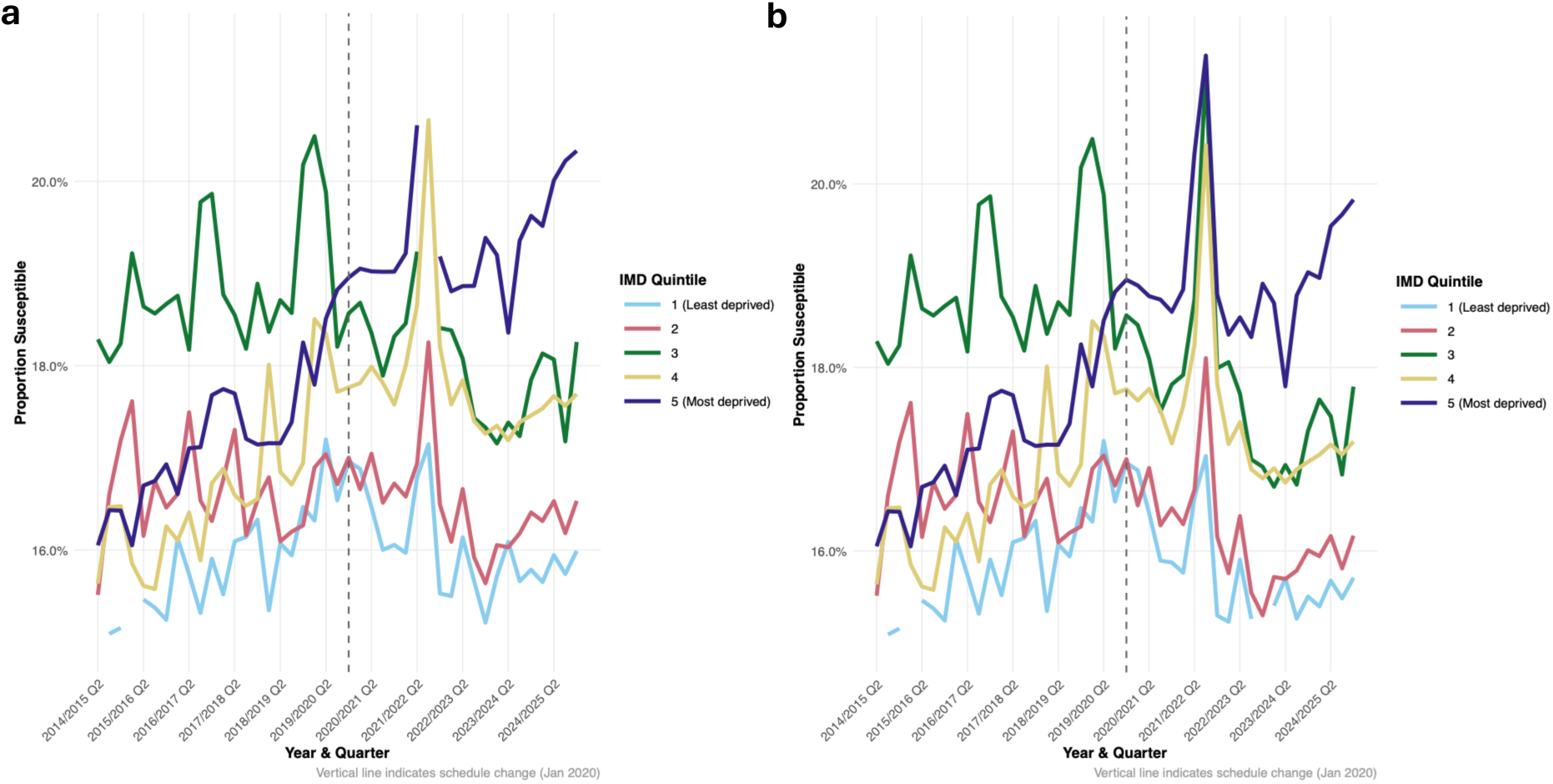
Estimated susceptibility to invasive pneumococcal disease by deprivation quintile: Baseline vs Alternate upper vaccine eNectiveness assumptions. **(a)** Baseline upper vaccine eXectiveness assumption. (b) Alternate upper vaccine eXectiveness assumption. Both scenarios show qualitative similar patterns to the estimates for the central vaccine eXectiveness assumptions. Quantitatively, for both the baseline and alternate scenarios the susceptibility estimates are reduced when applying the lower vaccine eXectiveness assumption compared to when applying the central vaccine eXectiveness assumption.

