## Supplementary material for "Inequalities in childhood pneumococcal conjugate vaccine uptake in England before and after the change from a 2+1 to 1+1 schedule: a longitudinal study": Tracked changes document.

##### 2.1.2. Linkage of vaccine uptake data to Index of Multiple Deprivation

Deleted: —an increase of 114% (95% CI 80–156%)

Deleted: model

Deleted: ,

Deleted: ing

Deleted: (

Deleted: for upper-tier local authorities (UTLAs) vaccine

Deleted: ,

Deleted: along with denominator eligible population counts for each cohort

Deleted: .

Deleted: ¶

The COVER data contained Office for National Statistics (ONS) UTLA codes. We therefore used the ONS UTLA codes to link the COVER vaccine uptake data to the 2019 Index of Multiple Deprivation (IMD) for England [15].

**Deleted:** ed

**Deleted:** via the Office for National Statistics (ONS) UTLA codes...

**Deleted:**

**Deleted:** We

**Deleted:** The City of London and

**Deleted:** .

**Deleted:** We cross-checked each cleaned quarterly COVER dataset against this validated list of 149 UTLAs. All COVER observations contained valid ONS local authority codes. However, not...

**Deleted:** 149

**Deleted:** upper-tier local authorities

**Deleted:** (12)

**Deleted:** IMD

**Deleted:** local authority

**Deleted:** ,

**Deleted:** f

**Deleted:** .

**Deleted:** W

**Deleted:** to reflect the population-weighted distribution of deprivation ...

between the mean 12-month primary coverage one year prior and the mean 24-month booster coverage at the reference quarter. As an example, to examine booster retention for Q1 2023 we used the mean 12-month primary coverage from Q1 2022 and the mean 24-month booster coverage from Q1 2023.

We stratified our analysis by schedule period: the pre-schedule change period (2013/2014 to 2019/2020 - the 2+1 schedule era) and the post-schedule change period (2020/2021 to 2024/2025 - the 1+1 schedule era). For each schedule period we identified statistical outliers using the interquartile range (IQR) method (i.e. outlier thresholds calculated as the 75th percentile plus 1.5 times the IQR).

#### 2.3. Pneumococcal disease susceptibility

**Deleted:** modelling

To estimate the vulnerability of specific birth cohorts to VT IPD at defined timepoints based on observed vaccine uptake patterns, we used a calculation combining the PCV coverage data with published vaccine effectiveness estimates for PCV13 against IPD. These estimates address susceptibility to VT IPD (i.e. IPD caused by serotypes covered by PCV13), but do not account for changes in risk form non-vaccine serotypes, which may vary over time.

**Deleted:** developed a simple protection model that

**Deleted:** ed

##### 2.3.1. The susceptibility calculation

**Deleted:** **model**

We classified as susceptible those at risk from contracting VT IPD due to either non-vaccination, partial vaccination (i.e. not completing the full schedule of doses) or the residual risk of disease after full vaccination [8,20].

Consequently, for each birth cohort (stratified by quarter) we calculated susceptibility as:

$$Susceptibility = p_0 + p_1(1-VE_1) + p_2(1-VE_2)$$

where $p_0$ corresponds to the proportion of children unvaccinated, $p_1$ the proportion who received only the primary doses (referred to as the ‘booster gap’ – the difference between the booster vaccine uptake at 24 months in the reference quarter and the primary vaccine uptake at 12 months one year prior) and $p_2$ the proportion who received the booster dose - for instances where booster vaccine uptake at 24 months in the reference quarter was greater than primary vaccine uptake at 12 months one year prior, we assumed none of the population had received primary doses only (i.e. $p_1 = 0$). $VE_1$ and $VE_2$ denote the estimated vaccine effectiveness against VT IPD after receiving the primary doses and booster dose, respectively.

We assumed that vaccine protection remained constant at the specified effectiveness levels throughout the observation period (up to 24 months post-booster). We acknowledge that our susceptibility calculation does not account for partial catch-up, waning immunity or indirect (herd) protection effects. Nonetheless, it does provide a first-order approximation of the proportion of each cohort that remains vulnerable to IPD based on coverage gaps and estimates of vaccine effectiveness against IPD, thereby allowing comparisons across deprivation quintiles and time periods.

**Deleted:** model

***Table 1. Vaccine effectiveness estimates against invasive pneumococcal disease.*** *We list the used lower, central and upper estimates. We sourced the estimates from European based studies reported by Savulescu et al.* [21].

| Dose | Schedule | Lower estimate | Central estimate | Upper estimate |
|---|---|---|---|---|
| Primary | 2+1 | 60 | 76.1 | 86 |
| | 1+1 (baseline assumption) | 29 | 60.6 | 78 |
| | 1+1 (alternate assumption) | 60 | 76.1 | 86 |
| Booster | All | 60 | 78.2 | 89 |

##### 2.3.2. Vaccine effectiveness assumptions

We selected vaccine effectiveness parameters for the susceptibility calculation from the European multi-country study by Savulescu et al. [21]. We expand below on our assumptions, with the vaccine effectiveness values used in the analysis summarised in Table 1.

**Deleted:** The study population included over 7 million eligible children across both 12-month cohorts (7,036,482 children) and 24-month cohorts (7,235,389 children).

**Formatted:** Font: +Body (Aptos)

#### 3.2. Trends and inequalities in PCV uptake

**Deleted:** The impact was more severe for booster doses, with all quintiles experiencing drops of 3-6 percentage points. ...

By 2023-2024, while 12-month coverage had largely recovered to pre-pandemic levels across most quintiles [Figure 1a], 24-month booster coverage remained depressed, particularly in more deprived areas [Figure 1b]. The deprivation gradient persisted, with quintiles 4 and 5 showing booster coverage 2-3 percentage points below the least deprived quintile - a gap that appears to have widened compared to the pre-2020 period.

As London's distinct population profile and scale can disproportionately influence national patterns, we generated equivalent results excluding London boroughs. The exclusion of London boroughs from our analysis substantially improved PCV uptake amongst quintile 3 UTLAs; London boroughs comprise 31% of quintile 3 areas but only 7-21% of other quintiles, creating a disproportionate impact on middle-deprivation coverage statistics. The exclusion of London boroughs had a more pronounced impact for booster uptake than primary dose uptake [Figure S4, S4 Text].

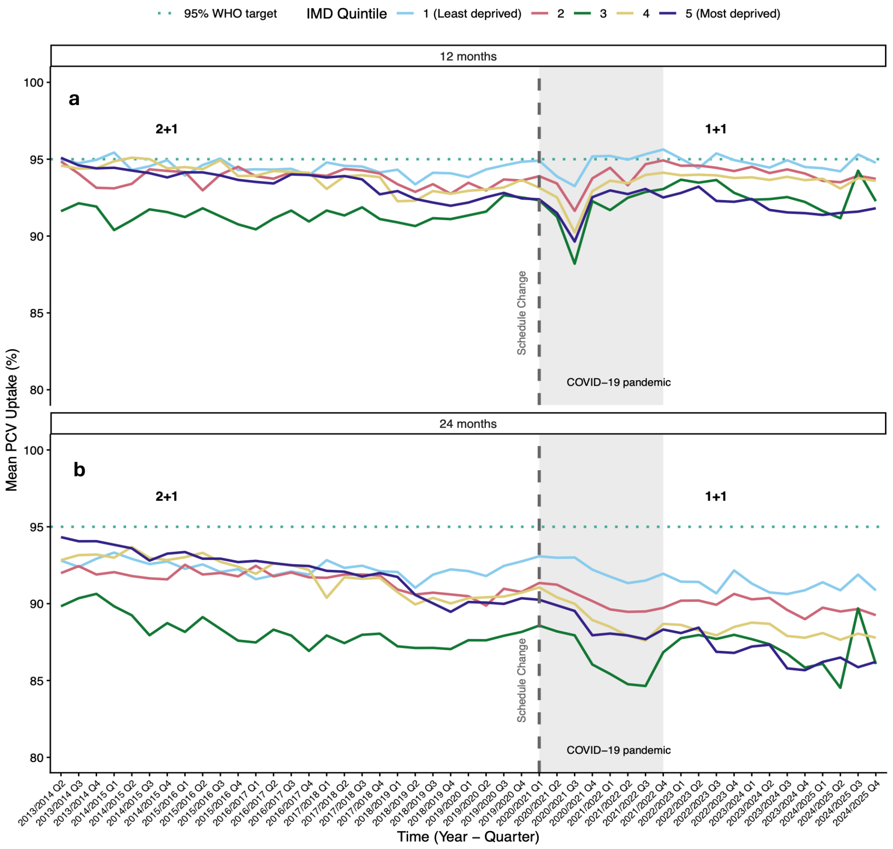


***Figure 1. Trends in Pneumococcal Conjugate Vaccine (PCV) uptake at 12 and 24 months by deprivation quintile, 2013–2024.*** We display for children in England, *grouped by deprivation quintile (IMD 1 = least deprived, IMD 5 = most deprived)*, the: **(a)** *average primary dose PCV uptake at 12 months*; **(b)** average *booster dose uptake at 24 months. The dotted* horizontal *line marks the WHO-recommended 95% target coverage. The dashed vertical line marks the January 2020 schedule change from a "2+1" to a "1+1" dosing regimen. The shaded grey area highlights the* COVID-19 pandemic period. We see a persistent and widening in vaccine uptake over time between the most and least deprived IMD quintiles.

**Deleted:** Excluding

**Deleted:** UTLAs

**Deleted:** coverage

**Deleted:** , demonstrating that this quintile's poor performance is driven by London-specific challenges rather than deprivation per se. ...

**Deleted:** Q

**Deleted:** The impact of London exclusion

**Deleted:** is

**Deleted:** coverage

**Deleted:** s

**Deleted:** ¶
¶

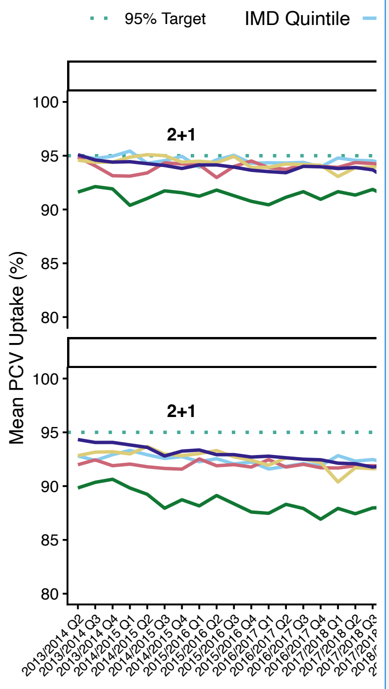


**Deleted:**

**3.2.2. Booster retention comparison: Deterioration in the retention of booster doses post the schedule change**

In the pre-schedule change period (2+1, 7-year average), Kensington and Chelsea, in London recorded the lowest overall coverage at 70.6% for the minimum of 12-month and 24-month uptake. The local authority with the largest booster gap was Hounslow, London (9.9%). We note that Lancashire recorded a 3.3% higher mean booster uptake by 24-months than mean primary dose coverage at 12-months in the pre-schedule change period. These data indicate potential reporting variations or catch-up vaccination patterns [Figure 2a]. The booster 'drop-off' distribution was heavily concentrated in the 1-2% range, with very few areas experiencing gaps exceeding 8 percentage points [Figure 2c].

In the post-schedule change period (1+1, 5-year average) the most concerning booster gaps were observed in Lewisham (11.3%), Greenwich (11.8%), and Brent (12.5%) - all London boroughs showing substantial deterioration in booster retention. Hackney recorded the lowest overall booster coverage at 65.5%, representing further decline in this already disadvantaged area [Figure 2b]. The post-schedule booster 'drop-off' distribution saw the modal bin move to approximately 2.5-3%, whilst becoming more dispersed and extending further into higher drop-off ranges. The maximum observed gap increased from approximately 9-10% to over 12% [Figure 2d].

These observations confirmed that the aggregate increase in mean booster gaps from 2.32% to 4.79% represented a systematic shift affecting many local authorities rather than being driven by a few extreme outliers. Deprivation-related patterns were also evident in both periods. Higher deprivation quintile areas (quintiles 4 and 5) more frequently appeared among the outliers with large booster gaps.

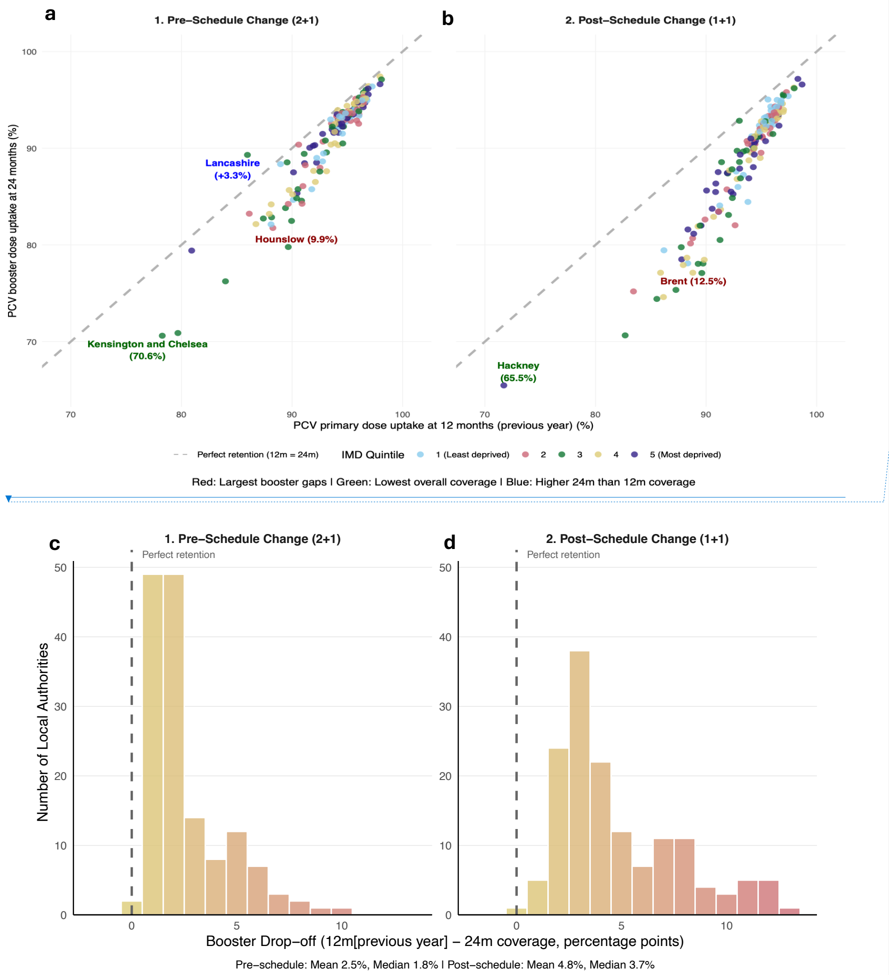


***Figure 2. PCV booster retention: Comparison before and after schedule change.*** ***(a,b)*** *Each dot represents a local authority. Points are coloured by deprivation quintile. We show average coverage during the:* ***(a)*** *2+1 schedule period (pre-2020, seven-year average);* ***(b)*** *1+1 schedule period (post-2020, five-year average). The dashed grey line shows perfect retention (equal uptake at 12 and 24 months). Dots below the line indicate booster drop-off. The post-schedule change period has many local authorities falling further below the diagonal, indicating a widespread deterioration in booster completion rates in England. Red labels signify the local authority with the largest booster gap before and after the schedule change (Pre-schedule change: Hounslow, 9.9%; Post-schedule change: Brent, 12.5%). Green labels correspond to the local authorities with the lowest overall coverage.* ***Blue*** *labels are local authorities with higher 24-month than 12-month coverage.* ***(c,d)*** *Distribution of booster drop-off during the:* ***(c)*** *2+1 schedule period (pre-2020, seven-year average);* ***(d)*** *1+1 schedule period (post-2020, five-year average). The colour gradient transitions from yellow (lower drop-off) to red (higher drop-off rates). The dashed vertical line at zero indicates perfect retention (equal 12-month and 24-month coverage). There is an evident rightward shift in the distribution toward higher drop-off rates in the post-schedule period.*

**Deleted:**

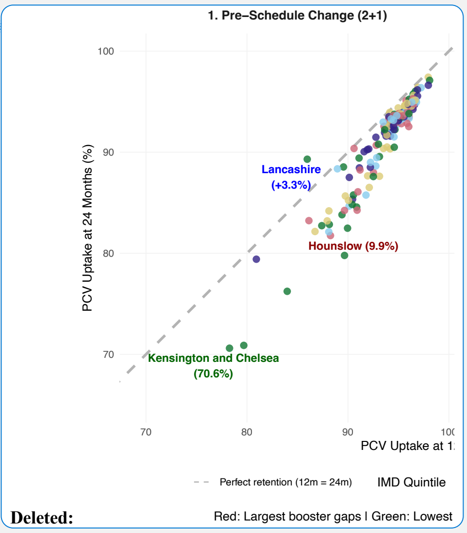

#### 3.3. Vaccine Type Invasive Pneumococcal Disease susceptibility

By combining observed vaccination uptake data with published vaccine effectiveness estimates, we estimated the proportion of children who remain susceptible to VT IPD across different time periods, deprivation levels, and geographic areas.

**Deleted:** modelled

**Deleted:** 1

**Deleted:** 7

**Deleted:** 8

**Deleted:** 0

**Deleted:** 1

**Deleted:** a

**Deleted:** The overall pattern shows more stable susceptibility levels under the 2+1 schedule compared to the increased variability observed during the 1+1 implementation period. Under the 2+1 schedule, susceptibility levels showed gradual increases from 2013 onwards, rising from approximately 30% to 31% by late 2019. The transition period marked by the dashed line (January 2020) coincided with a sharp spike in susceptibility, reaching 49% during 2020-2021 Q1 [Figure 3b]. ...

**Deleted:** ¶

**Deleted:** , left panel

**Deleted:** , right panel

**Deleted:** for the 1+1 schedule

**Deleted:**

**Deleted:** similar spatial distributions of susceptibility levels and ...

**Deleted:** 5

estimates across IMD quintiles were between 25-31%), the susceptibility estimates were elevated when applying the lower vaccine effectiveness assumption (between 42-48%; Figure S8, S6 Text) and reduced when applying the higher vaccine effectiveness assumption (between 15-22%; Figure S9, S6 Text).

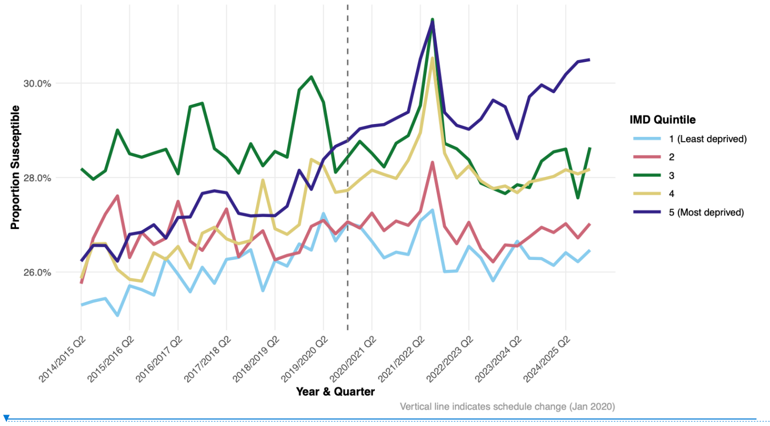


***Figure 3. Estimated susceptibility to invasive pneumococcal disease by deprivation quintile: Baseline vaccine effectiveness assumption.*** *Lines represent IMD quintiles (1 = least deprived, 5 = most deprived). The vertical dashed line marks the January 2020 schedule change. We observe persistent deprivation gradients and elevated vulnerability in quintiles 3 and 5 throughout the study period.*

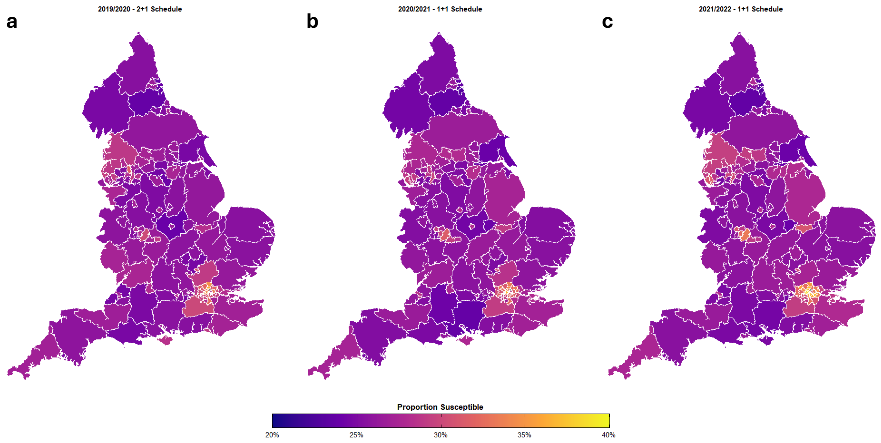


***Figure 4. Geographic variation in estimated susceptibility to invasive pneumococcal disease across upper-tier local authorities in England.*** *We show average susceptibility by upper-tier local authority for: (a) 2019/2020 (2+1 schedule); (b) 2020/2021 (transition); (c) 2021/2022 (1+1 schedule). Colour scale: purple (20% susceptibility) to yellow (35% susceptibility). Estimates based on observed coverage and baseline vaccine effectiveness assumptions (1+1 primary VE = 60.6%). Geographic patterns remain relatively consistent across periods, with most areas showing 20-30% susceptibility. Pockets of higher vulnerability are visible in urban centres, reflecting local variation in vaccination coverage and population characteristics. For the period 2021/2022 we have filled the 'Northamptonshire' UTLA based on a weighted average from the 'West Northamptonshire' & 'North Northamptonshire' data.*

Deleted: Page Break ¶

Deleted:

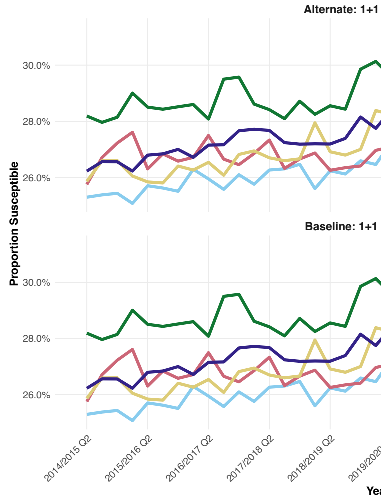


Formatted: Normal

Deleted: *S*

Deleted: *I*

Deleted: *P*

Deleted: *D*

Deleted: *D*

Deleted: *Q*

Deleted: *vs Alternate VE*

Deleted: *A*

Deleted: *. We display **the upper panel**: alternate assumption (1+1 primary dose VE = 76.1%); lower panel: baseline assumption (1+1 primary dose VE = 60.6%).*

Deleted: *Both scenarios show similar susceptibility patterns over time, with*

Deleted: *,*

Deleted: *,*

Deleted: *and*

Deleted: *panel*

### 4. Discussion

This study examined trends and inequalities in PCV13 uptake across England following the January 2020 transition from a 2+1 to 1+1 dose regimen. The analysis revealed three critical findings: booster retention deteriorated during the 1+1 schedule; socioeconomic inequalities in vaccine uptake persisted and widened (particularly affecting the most deprived communities); a growing population-level and inequitable susceptibility to VT IPD.

EMH is affiliated to the NIHR Health Protection Research Unit in Emerging and Zoonotic Infections (NIHR207393). EMH is funded by The Pandemic Institute, formed of seven founding partners: The University of Liverpool, Liverpool School of Tropical Medicine, Liverpool John Moores University, Liverpool City Council, Liverpool City Region Combined Authority, Liverpool University Hospital Foundation Trust, and Knowledge Quarter Liverpool (EMH is based at The University of Liverpool). The views expressed are those of the author(s) and not necessarily those of NIHR, the Department of Health and Social Care or The Pandemic Institute.

The funders had no role in study design, data collection and analysis, decision to publish, or preparation of the manuscript. For the purpose of open access, the authors have applied a Creative Commons Attribution (CC BY) licence to any Author Accepted Manuscript version arising from this submission.

#### Data and code availability statement

All data utilised in this study are publicly available. We state relevant references and data repositories stated within the main manuscript.

Data and code associated with the study are available in the following repository: https://github.com/aaliswalker/DASC500-Dissertation---Inequalities-in-Childhood-Pneumococcal-Vaccine-Uptake-in-England

Archived code: https://doi.org/10.5281/zenodo.18420636

**Deleted:** https://doi.org/10.5281/zenodo.17854535

#### Declaration of interests

DH and NF are currently in receipt of grant support from Seqirus UK for the evaluation of influenza vaccines in the UK; NF is in receipt of funding from GSK in relation to malaria vaccines; DH has also received grants from Sanofi Pasteur, and Merck and Co (Kenilworth, NJ) for rotavirus strain surveillance, received honorariums for presentation at a Merck Sharp and Dohme (UK) symposium on vaccines and has consulted on rotavirus strain surveillance; PI and EMH have nothing to disclose.

### Supplementary Material

#### S1 Text. COVER data preparation

We downloaded all pneumococcal vaccine uptake data from the COVER programme for the years 2013 to 2024, spanning a total of 52 quarterly files. These files varied in structure, format (.xlsx, .ods), and naming conventions. To ensure consistency, we renamed all files using a uniform format: 20XX_QX for pre-2020 files and 20XX QX for post-2020 files.

We imported all quarterly PCV uptake files into R and combined them into a single dataset for analysis. Due to inconsistencies across years—including changes in sheet names, column headers, and data formatting—we wrote custom cleaning scripts to extract only the relevant PCV tables (i.e., 12-month and 24-month PCV coverage and population denominators). We filtered out unrelated tables (e.g. MMR, Hib/MenC), and harmonised all extracted columns using a consistent naming convention (e.g., PCV_12m, PCV_24m, ONS_Code). [Table S1]

To prevent data type mismatches, we initially read all columns as character vectors, before converting them to appropriate types for analysis (e.g., numeric for coverage percentages and denominators, factor for categorical variables). To ensure that we handled missing values consistently, we recoded any value starting with 'N' (e.g., "N.A.") or containing extraneous symbols (such as those beginning with '[' e.g [z]) as NA.

***Table S1. Variables used for data preparation.***

| Variables | Meaning |
|---|---|
| ONS_Code | ONS Upper Tier Local Authority code (geographic unit of analysis) |
| PCV_12m | Percentage uptake of the pneumococcal vaccine at 12 months |
| PCV_24m | Percentage uptake of the booster dose (at 24 months) |
| Population_12m | Eligible population denominator for 12-month coverage |
| Population_24m | Eligible population denominator for 24-month coverage |
| Year | Year the data relates to (e.g. "2019/2020") |
| Quarter | Quarter of the data year (e.g. Q1, Q2, Q3, Q4) |
| Timepoint | 0 = Q1 2013, 1 = Q2 2013, etc. |
| Vaccine_Schedule | Binary indicator: 0 = 2+1 schedule (pre-2020), 1 = 1+1 schedule (2020+) |

NB: As documented on the COVER data, there are a variety of reasons why some entries in the original data files are censored, including but not limited to:

a. Small denominators, with data censored to protect the identities of patients
b. Data quality issues
c. Data missing or unavailable

**Deleted:** ¶ ... [1]

**Formatted:** Font: +Body (Aptos), 20 pt

**Formatted:** Font: +Body (Aptos)

d. Unrecognisable code from EMIS: These data are collected through routine reporting by Child Health Information Systems (local NHS digital systems that track childhood immunizations and health records, including EMIS - Egton Medical Information Systems, which is one of the main GP IT systems used across England; for more details, visit https://digital.nhs.uk/services/gp-it-futures-systems/im1-pairing-integration/emis-pfs-suppliers).

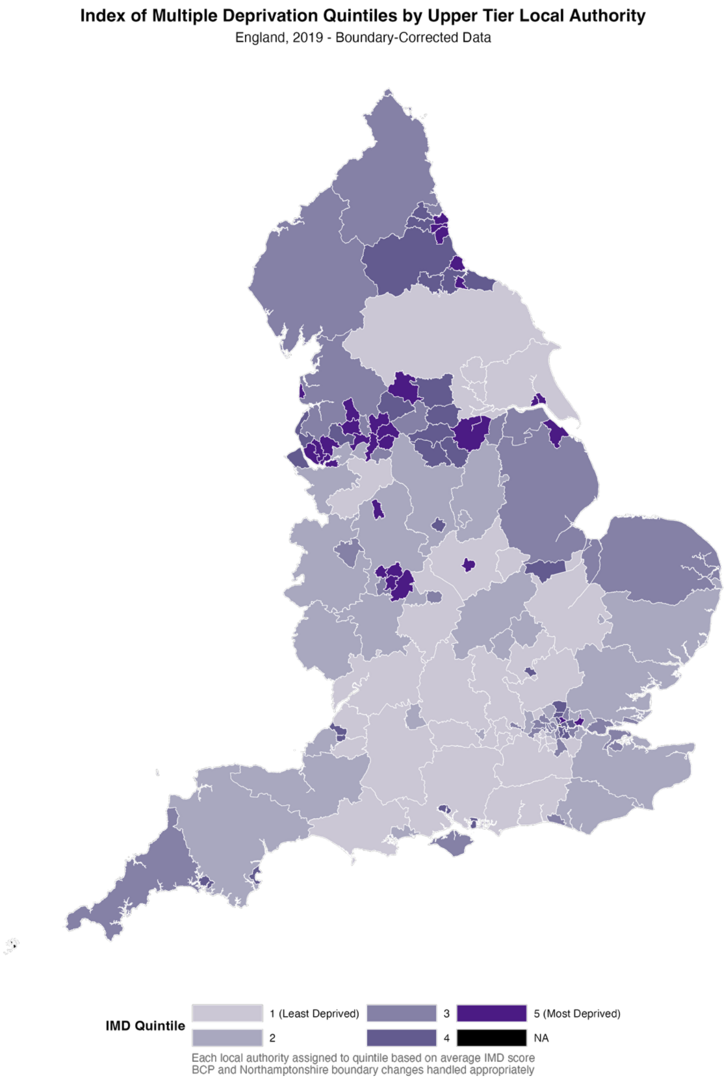


**Figure S1. Assignment of UTLAs in England to IMD quintiles.** *These classifications used UTLA-level Index of Multiple Deprivation (IMD) summary data from the English Indices of Deprivation 2019* [15]*. We assigned each UTLA to an IMD quintile based on the average IMD score for each UTLA. Shading denotes the IMD quintile associated with that UTLA, with the lightest shading representing the least deprived UTLAs (quintile 1) and the darkest shading representing the most deprived UTLAs (quintile 5). We assigned as NA the UTLAs not retained for our study (Isle of Scilly, The City of London).*

#### S4 Text. Additional PCV uptake figures

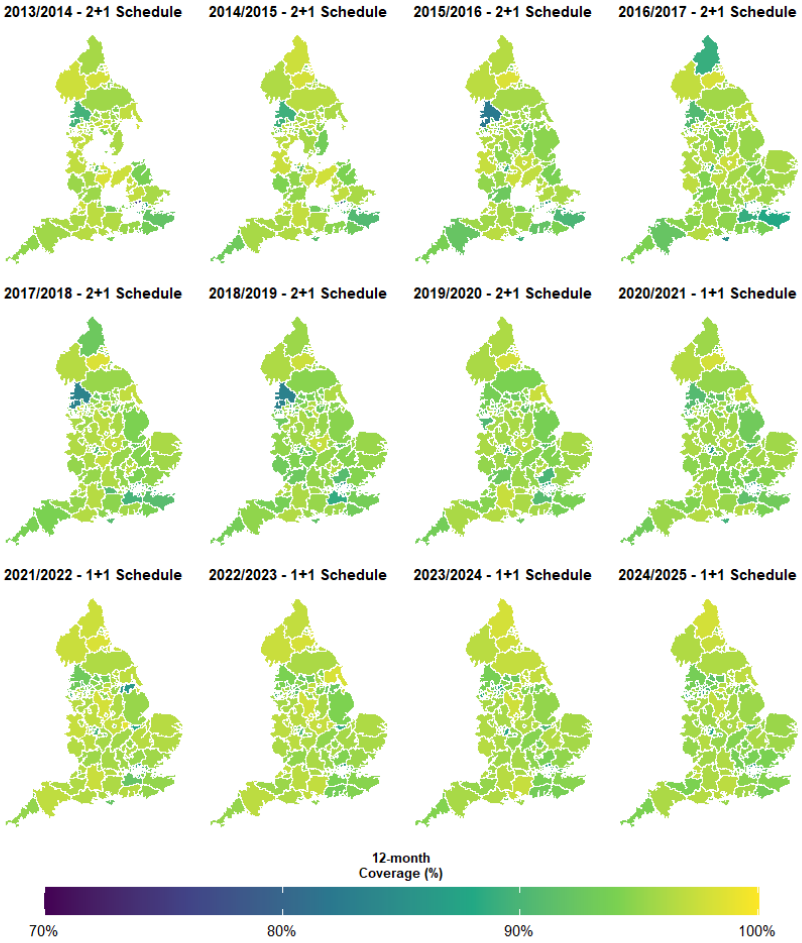


***Figure S2. Geographic variation in PCV primary coverage (12 months) across local authorities in England, 2013/2014 to 2024/2025.*** *The colour scale ranges from dark blue/purple (70% coverage) to bright green/yellow (100% coverage), so brighter areas indicate higher vaccination rates while darker areas show lower uptake. The maps are arranged chronologically from left to right, top to bottom, spanning both the 2+1 schedule period (2013-2020), and the 1+1 schedule period (2021-2025).*

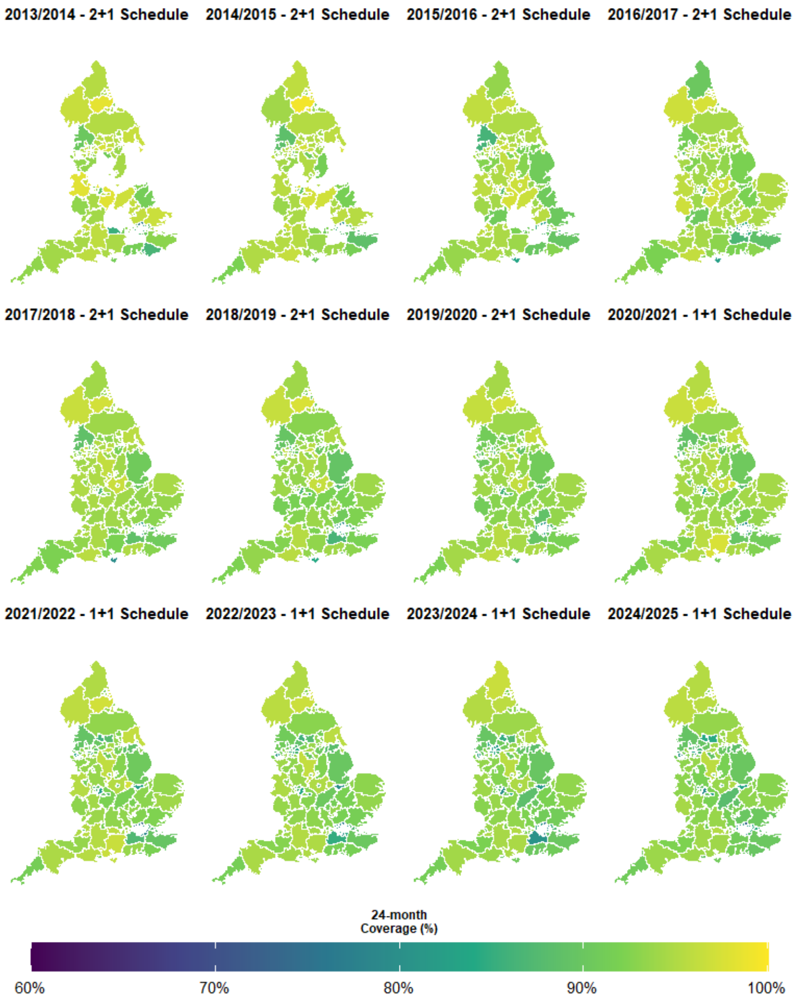


***Figure S3. Geographic variation in PCV booster coverage (24 months) across local authorities in England, 2013/2014 to 2024/2025.*** *The colour scale ranges from dark blue/purple (60% coverage) to bright yellow (100% coverage), where brighter green/yellow areas indicate higher booster uptake and darker blue areas show lower coverage. The maps are arranged chronologically from left to right, top to bottom, spanning both the 2+1 schedule period (2013-2020), and the 1+1 schedule period (2021-2025).*

**a**

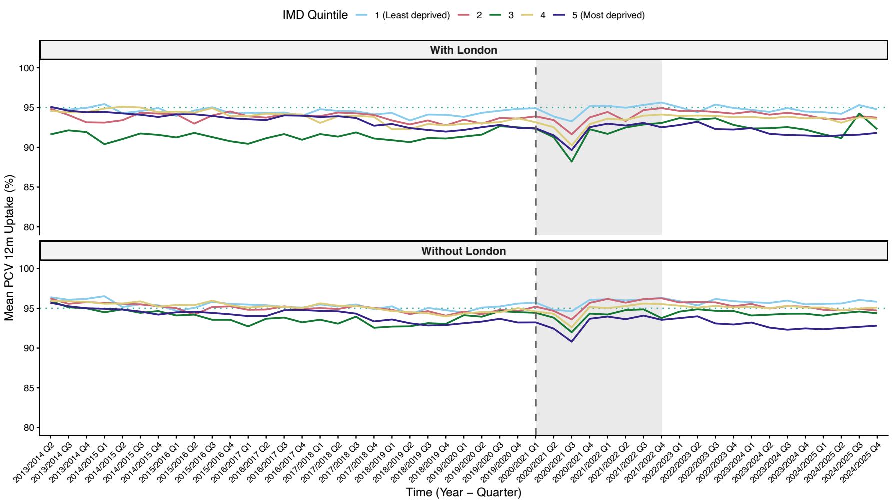


**b**

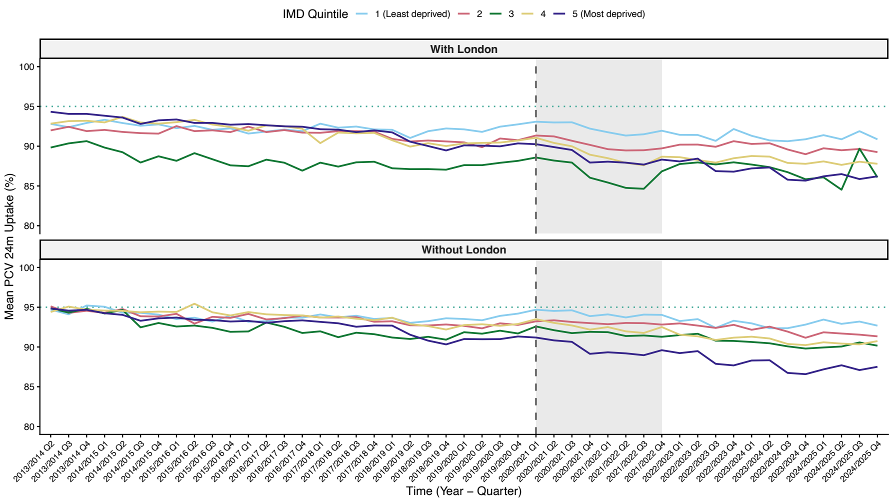


**Figure S4. Impact of London on PCV Coverage by deprivation quintile.** *Comparison of uptake trends for: **(a)** primary doses at 12 months; **(b)** booster dose at 24 months. In each panel, the upper plot includes 32 London boroughs (i.e. all London boroughs except the City of London), whereas the lower panel does not include any London boroughs. Lines represent IMD quintiles (1 = least deprived, 5 = most deprived). Dashed vertical line: schedule change (January 2020); shaded area: COVID-19 period. The impact of London exclusion is more pronounced for booster coverage than primary doses.*

**Deleted:** D

**Deleted:** Q

**Deleted:** *at*

**Deleted:** *'s 76*

**Deleted:**

**Deleted:** *'s 32*

### S5 Text. Cumulative VT IPD susceptibility estimates indicate widening socioeconomic inequalities

To better understand the growing burden of disease risk over time, we calculated the cumulative number of susceptible children by birth cohort (quarterly). This analysis demonstrates how even small reductions in uptake can lead to a steady accumulation of susceptible individuals, which may have implications for herd protection and future disease resurgence if left unaddressed [Figure S5a]. Susceptibility patterns revealed persistent and widening inequalities by deprivation level. Although overall susceptibility levels appeared similar before and after the schedule change, the more deprived areas (quintiles 3 and 5) experienced disproportionately higher vulnerability throughout the study period [Figure S5b].

**Deleted:** Geographic patterns remained consistent across both VE assumptions, with similar spatial distributions of susceptibility levels and no dramatic changes in regional vulnerability patterns [Figure S6].¶

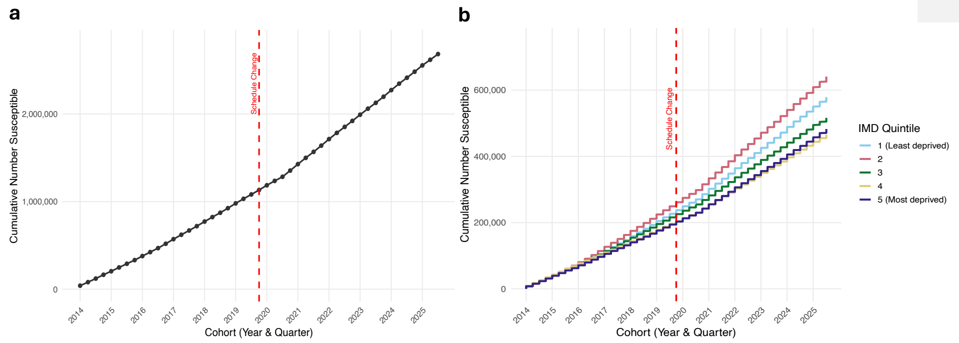


***Figure S5. Cumulative number of susceptible children by quarter.*** *We calculated susceptibility on observed uptake and literature-based vaccine effectiveness.* ***(a)*** *Each point represents the total number of children estimated to remain susceptible to invasive pneumococcal disease (IPD) after vaccination, cumulatively summed from 2013/2014 Q2 to 2024/2025 Q3.* ***(b)*** *Stratification by deprivation quintiles. (1 = least deprived, 5 = most deprived). The x-axis tracks cohort entry by year and quarter.*

**Deleted:** *Susceptibility*

**Deleted:** *is modelled based*

#### S6 Text. Findings robust to alternate vaccine effectiveness assumptions

When applying the alternate central vaccine effectiveness assumption in our susceptibility calculation, throughout most of the study period, we found quintiles 3 and 5 (most deprived) had the highest susceptibility levels, while quintile 1 (least deprived) generally maintained the lowest susceptibility [Figure S6]. Geographic patterns remained consistent across both VE assumptions, with similar spatial distributions of susceptibility levels and no dramatic changes in regional vulnerability patterns [Figure S7].

When applying either the lower or upper vaccine effectiveness estimates, rather than the central vaccine effectiveness estimates [Table 1], we found qualitative patterns were maintained [Figures S8 & S9]. Quantitatively, comparing susceptibility estimates to when applying the central vaccine effectiveness assumption (where susceptibility estimates across IMD quintiles were between 25-31%), the susceptibility estimates were elevated when applying the lower vaccine effectiveness assumption (between 42-48%; Figure S8) and reduced when applying the higher vaccine effectiveness assumption (between 15-22%; Figure S9).

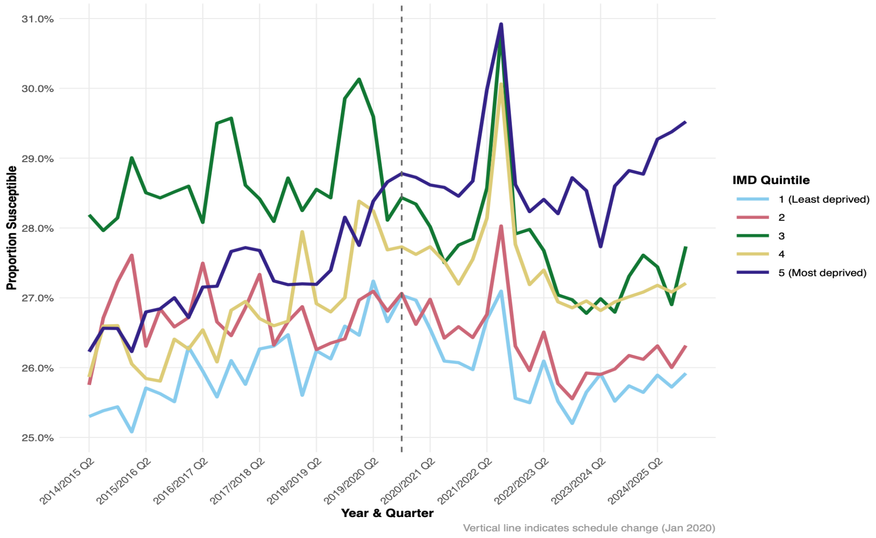


**Figure S6. Estimated susceptibility to invasive pneumococcal disease by deprivation quintile: Alternate central vaccine effectiveness assumption** (1+1 primary dose VE = 76.1%). Lines represent IMD quintiles (1 = least deprived, 5 = most deprived). The vertical dashed line marks the January 2020 schedule change. We observe persistent deprivation gradients and elevated vulnerability in quintiles 3 and 5 throughout the study period.

**Formatted:** Centred

**Formatted:** Font: Bold

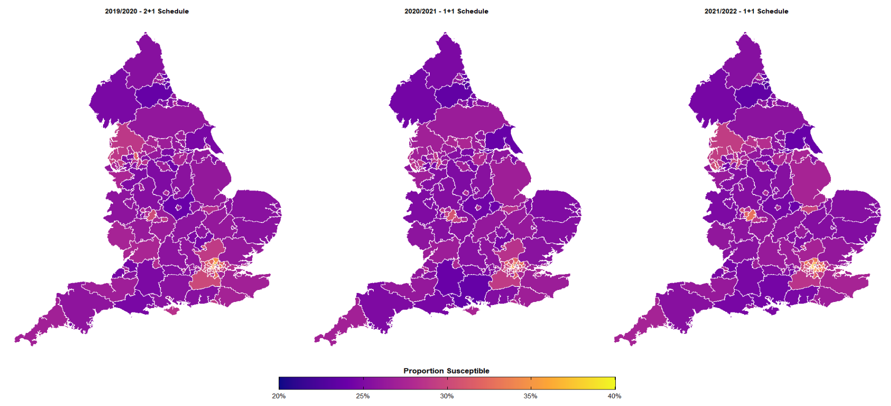


**Figure S7. Geographic variation in IPD susceptibility: Alternate central vaccine effectiveness assumption.** *We show average susceptibility by upper-tier local authority for 2019/2020 (2+1 schedule), 2020/2021 (transition), and 2021/2022 (1+1 schedule). Colour scale: purple (20% susceptibility) to yellow (35% susceptibility). Estimates based on alternate vaccine effectiveness assumption (1+1 primary VE = 76.1%, matching completed 2+1 primary course with booster 78.2%). Most areas show 20-30% susceptibility, with persistent pockets of higher vulnerability in urban centres.*

**Formatted:** Font: 11 pt, Bold

**Formatted:** List Paragraph

**Deleted:** 6

**Deleted:** V

**Deleted:** S

**Deleted:** A

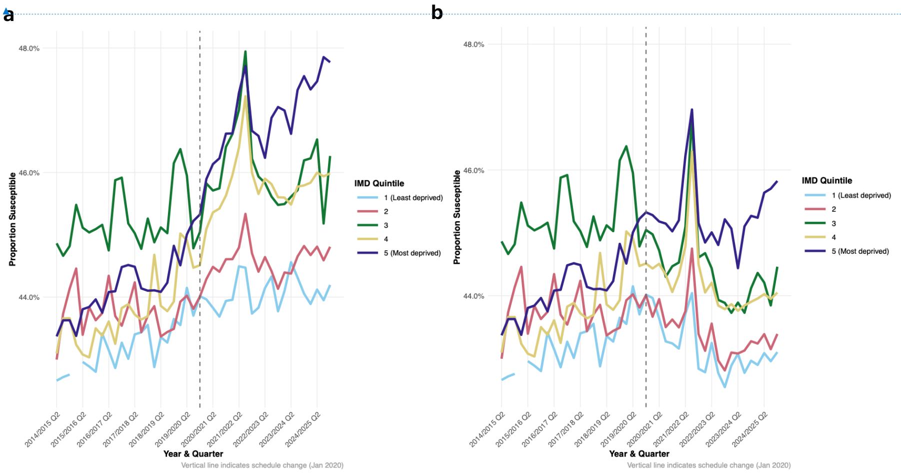


**Figure S8. Estimated susceptibility to invasive pneumococcal disease by deprivation quintile: Baseline vs Alternate lower vaccine effectiveness assumptions**. ***(a)*** *Baseline lower vaccine effectiveness assumption.* ***(b)*** *Alternate lower vaccine effectiveness assumption. Both scenarios show qualitative similar patterns to the estimates for the central vaccine effectiveness assumptions. Quantitatively, for both the baseline and alternate scenarios the susceptibility estimates are elevated when applying the lower vaccine effectiveness assumption compared to when applying the central vaccine effectiveness assumption.*

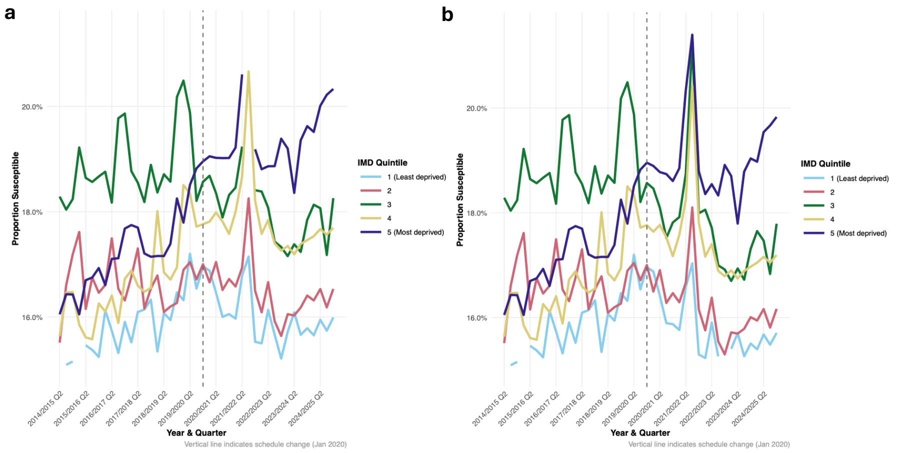


**Figure S9. Estimated susceptibility to invasive pneumococcal disease by deprivation quintile: Baseline vs Alternate upper vaccine effectiveness assumptions**. **(a)** Baseline upper vaccine *effectiveness assumption.* ***(b)*** *Alternate upper vaccine effectiveness assumption. Both scenarios show qualitative similar patterns to the estimates for the central vaccine effectiveness assumptions. Quantitatively, for both the baseline and alternate scenarios the susceptibility estimates are reduced when applying the lower vaccine effectiveness assumption compared to when applying the central vaccine effectiveness assumption.*

**Formatted:** Font: +Body (Aptos), 9 pt, Not Bold, Italic

**Formatted:** Font: Not Italic

**Formatted:** Font: Not Bold, Not Italic

**Formatted:** Heading 4

**Formatted:** Font: Not Bold

**Formatted:** Font: +Body (Aptos), 9 pt, Not Bold, Font colour: Text 1

**Formatted:** Font: +Body (Aptos), 9 pt, Font colour: Text 1

| Page 21: [1] Deleted | Hill, Edward [edmhill] | 25/01/2026 16:05:00 |
| --- | --- | --- |
